# Clinically-feasible white matter fiber tractography in peritumoral zones with cerebral vasogenic edema

**DOI:** 10.1101/2025.09.12.25335440

**Authors:** Patryk Filipiak, Timothy M. Shepherd, Kamri Clarke, Dimitris G. Placantonakis, Fernando E. Boada, Steven H. Baete

## Abstract

In diffusion MRI, vasogenic edema manifests as a major fraction of isotropic water that dilutes the anisotropic intraaxonal portion of the signal. Many tractography algorithms mistake vasogenic edema for the white matter boundary and terminate tracking to prevent producing spurious streamlines. As a result, visual representations of fascicles traversing edema are often compromised, limiting the clinical utility of tractography in neurosurgery.

We address this hurdle with ODF-Fingerprinting (ODF-FP) — a dictionary-based fiber reconstruction algorithm that accommodates variability of neural tissue. By adding a regularization term to the ODF-FP matching formula, we counterbalance the drop of diffusion anisotropy in edematous regions to improve white matter fiber identification.

We test our approach in 19 glioma cases with significant peritumoral vasogenic edema. For each case, we quantify the volume of the reconstructed white matter tracts immersed in edema, then we use the cortical regions activated during task-based functional MRI as validation for tractography. To assess the potential for clinical translation, we additionally test the performance of our reconstruction method on subsampled single-shell diffusion-weighted images, which contemporary clinical scanners can acquire within a few minutes.

## 1. Introduction

Cerebral edema, or swelling of the brain, is a major confounding factor for diffusion MRI (dMRI) tractography [1]. Excessive fluids accumulated in edematous White Matter (WM) decrease anisotropy of water self-diffusion inside neural tissue, which reflects in premature termination of tracked fascicles [2]. Therefore, visual representations of WM fibers traversing edema are often compromised, limiting the clinical utility of tractography in neurosurgery [2, 3, 4].

We addressed this hurdle by proposing a modification of the dictionary-based WM fiber reconstruction technique called Orientation Distribution Function Fingerprinting (ODF-FP) [5, 6] to accommodate the abundance of fluids in the impacted tissue. We tested our approach in 19 glioma cases with significant peritumoral vasogenic edema. For each case, we quantified the volume of the reconstructed WM tracts immersed in edema, then we used the cortical regions activated during task-based functional MRI (fMRI) as validation for tractography.

### 1.1. Vasogenic edema in glioma

Buildup of liquids in cerebral edema is typically caused by tissue injuries [7], such as stroke, trauma, or tumor. In the latter case, when tumor infiltration disrupts blood-brain barrier, brain capillaries are leaking fluids that accumulate in the extra-axonal space [8]. This type of edema is called *vasogenic*, which means “originating from blood vessels”.

The volume of vasogenic edema caused by aggressive brain tumors, such as high-grade glioma, often surpasses the tumor volume itself [9]. Consequently, MRI abnormalities are visible in a vast portion of peritumoral tissue, especially in Fluid-Attenuated Inversion Recovery (FLAIR) T2-weighted images. Displacement and damage to WM tracts caused by peritumoral edema can lead to development of cognitive impairments in about 65% of patients [10]. Despite these abnormalities, edematous WM often retains a part of its eloquent functions [7]. Therefore, surgical excision of the impacted tissue is likely to cause neurological deficits [11].

For maximally safe tumor resection, neurosurgeons need to reconcile two conflicting criteria: On the one hand, maximizing the resection volume can prolong the estimated progression-free survival by 61-90 months [12]. On the other hand, safeguarding eloquent WM is necessary to ascertain quality of life without postoperative deficits [11]. Under these circumstances, the ability to identify and visualize eloquent peritumoral fiber structures, which remain functional despite edema, is critical to inform neurosurgeons about the WM regions that must be spared.

### 1.2. Impact of vasogenic edema on dMRI tractography

In dMRI, vasogenic edema manifests as a major fraction of isotropic water that dilutes the anisotropic intraaxonal portion of the signal [1]. Many tractography algorithms mistake vasogenic edema for the WM boundary — since both cause a sudden drop in Fractional Anisotropy (FA) — so they terminate tracking to prevent producing streamlines outside WM [4, 13]. As a result, fibers traversing vasogenic edema are underrepresented in tractography-based visualizations, even if they remain functional [1].

To address this issue, Pasternak et al. [14, 15] proposed Free Water Elimination Diffusion Tensor Imaging (FWE-DTI) technique, which decomposes the diffusion-weighted signal into two compartments — an isotropic component representing edema and a residual diffusion tensor, FWE-DT, representing neural tissue. By computing FA maps from FWE-DT, rather than from the diffusion tensor of the original signal, the authors attempted to compensate the anisotropy drop due to vasogenic edema, and thus reinforce tracking. However, this conceptual shift from DTI to the bicompartmental FWE-DTI has increased the complexity of the signal representation, which no longer provided a unique parametrization of clinically prevalent singleshell dMRI.

Freewater EstimatoR using iNtErpolated iniTialization (FERNET) [16] mitigated the non-uniqueness problem by using the combination of gradient descent algorithm and the initialization strategy that incorporated the b0-image (i.e., the signal without diffusion weighting) to support convergence toward biologically plausible values. This modification helped to stabilize the FWE-DTI parameter fitting in edematous brain on a single-shell data, although the major limitation of the technique remained in the underlying diffusion tensor model [17], which is unable to reconstruct complex fiber structures [13].

Recently, Hakhu et al. [18] showed that Neurite Orientation Dispersion and Density Imaging (NODDI) [19] outperforms DTI-based approaches in separating the free-water content of edematous zones surrounding meningiomas. However, the proposed method — similarly to FWE-DTI and FERNET — is unable to represent crossing fibers, which are ubiquitous in the brain [20].

Despite the demonstrated impact of vasogenic edema on dMRI tractography, neither of the existing edema compensation mechanisms is used in everyday practice due limited clinical research [21] and difficulty to estimate the free-water content reliably [22]. We aim to fill these gaps by proposing a clinically-feasible WM fiber reconstruction method based on ODF-FP described below. Note that our approach bypasses the need to explicitly estimate and extract the free-water content in edema, which is the bottleneck of the methods introduced earlier.

### 1.3. Orientation distribution function fingerprinting

Fingerprinting is a computational technique to efficiently compare large data objects using their simplified representations, called fingerprints [23]. ODF-FP employs this technique to reconstruct WM fibers based on their ODFs. For this, ODF-FP maintains an extensive dictionary of synthetically generated ODFs and matches them with the ODFs calculated from dMRI to look up the underlying WM fiber structure [5, 6] (Figure 1).

**Figure 1.**
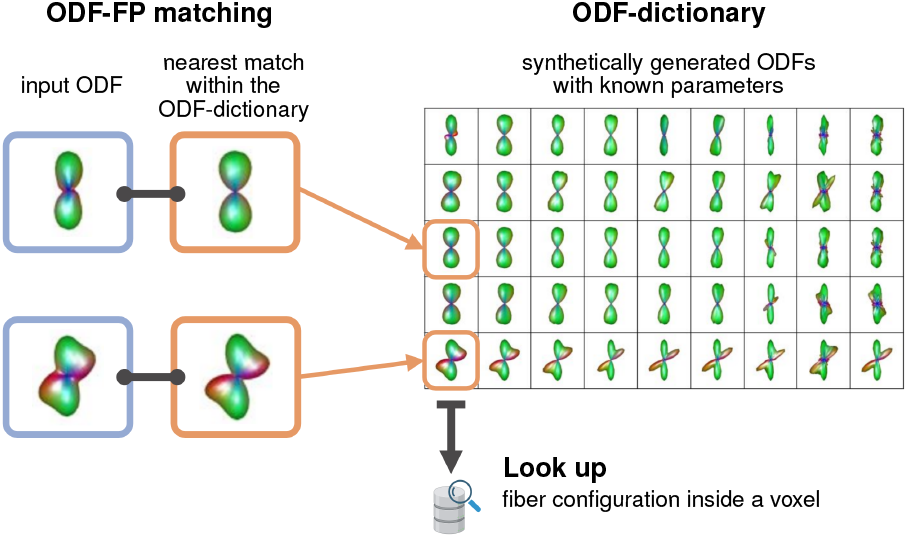
Schematic workflow of ODF-FP: Input ODFs, computed from dMRI, are matched with their most similar counterparts within an ODF-dictionary. The dictionary contains synthetically generated ODFs with known microstructure parameters, which can be looked up to reconstruct the underlying fiber structure.

Many standard methods, such as Generalized Qsampling Imaging (GQI) [24] or Constrained Spherical Deconvolution (CSD) [25], reconstruct fiber directions by finding local maxima of ODFs, called ODF peaks. This standard approach, although simple and generally effective [13], has limited robustness to signal noise [26], which leads to false positives (due to spurious peaks) or false negatives (due to blurred peaks). ODF-FP alleviates this issue by replacing the peakfinding technique with pattern matching, for which it uses ODFs as fiber structure fingerprints [5]. As a result, ODF-FP can extract information embedded in entire ODF shape, rather than solely localize its peaks. In the case of edema, ODF-FP does not need to explicitly separate the free-water content. Instead, it matches the fingerprints of edematous regions with the corresponding fingeprints in a sythetically generated ODFdictionary.

The efficacy of ODF-FP was demonstrated in tracking major WM fascicles, including pyramidal tracts [6] and optic pathways [27], where complex fiber structures contributed to uncertainties in peak finding. In this paper, we employ ODF-FP to address the problem of WM fiber identification in edematous tissue. To further improve the efficacy of our approach, we introduce a diffusion anisotropy boosting mechanism that counterbalances the drop of anisotropy caused by vasogenic edema.

## 2. Theory

Define **ODF-dictionary** *𝒟* as a finite set of synthetically generated normalized ODFs [6]:

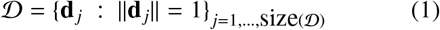

where each element **d** *_j_* ∈ *𝒟* represents a configuration of *N_j_* ≥ 0 fibers, particularly:

- *N_j_* = 0 (free water) or
- *N_j_* = 1 (single fiber) or
- *N_j_* ≥ 2 (crossing fibers).

Let **x** be a normalized ODF (i.e., ∥**x**∥ = 1) calculated from dMRI. To reconstruct fiber directions, ODF-FP finds the element:

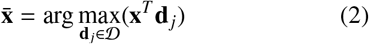

that maximizes the cosine similarity to **x**. However, dMRI signal distortions embedded in **x** pose a risk of overfitting to noise. Therefore, we use the **regularized ODF-FP matching formula**:

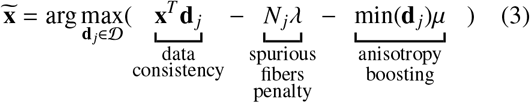

for *λ, µ* ≥ 0.

The first regularization term, introduced earlier [6], eliminates spurious fibers by promoting dictionary elements with lower *N_j_*.

In this work, we propose the second regularization term to **boost di**ff**usion anisotropy** by penalizing dictionary elements with high global minima, as illustrated in Figure 2. Note that ODFs with narrow peaks, desired for accurate tractography, are characterized by lower min(**d** *_j_*) than the ones perturbed by noise, e.g., due to edema. To mitigate this process, we promote ODF-dictionary elements with lower global minima, using the adjustable factor µ. Particularly, increased µ_edema_ in edematous WM can prevent premature termination of fiber tracking, as we will demonstrate later.

**Figure 2.**
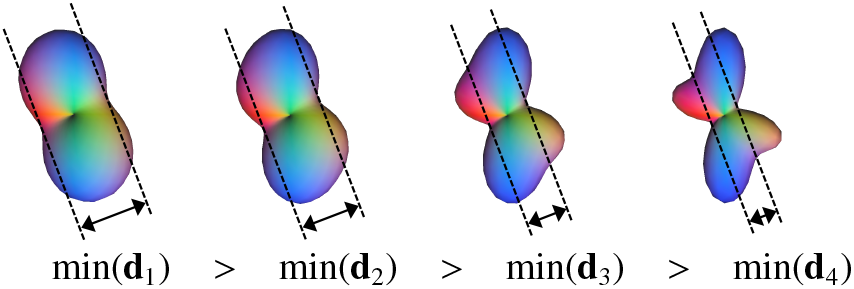
Two crossing fibers represented by the normalized ODFs **d**_1_, **d**_2_, **d**_3_, **d**_4_ with different global minima. Notably, the lowest minimum, i.e., min(**d**_4_), ensures the most distinguishable peaks, which is desired for accurate tractography.

## 3. Methods

This retrospective study was approved by the Internal Review Board (IRB) of NYU Langone Health (protocol number i18-00124). Access to previously collected anonymized data did not require additional consent from the participants. All images were acquired with a clinical 3T Siemens Prisma (Erlangen, Germany) MR scanner.

### Patients

We considered all patients scheduled for surgical resection of glioma who underwent dMRI and fMRI acquisition during the same imaging session at NYU Langone Health between January 1, 2023 and December 31, 2024. Among those, we selected all cases with peritumoral vasogenic edema infiltrating at least one of the major WM fascicles, i.e., Arcuate Fasciculus (AF), Superior Longitudinal Fasciculus III (SLF3), Inferior Fronto-Occipital Fasciculus (IFOF), Frontal Aslant Tract (FAT), Corticospinal Tract (CST), or Corticobulbar Tract (CBT). We then eliminated the cases were dMRI or fMRI were compromised by significant artifacts. Thus obtained data set contained preoperative MRI of 19 patients (Table 1): 6 females /13 males, aged 32–73 y/o (mean ± std: 53 ± 13 y/o).

**Table 1:** Summary of patients data including the tumor type, WHO Grade (2, 3, 4, or undefined), tumor location in the Left or Right hemisphere (L/R). The ipsilateral fascicles traversing vasogenic peritumoral edema and the corresponding fMRI tasks used as reference are marked with a full circle (•). *Fascicle names:* AF – Arcuate Fasciculus, SLF3 – Superior Longitudinal Fasciculus III, IFOF – Inferior Fronto-Occipital Fasciculus, FAT – Frontal Aslant Tract, CST – Corticospinal Tract, CBT – Corticobulbar Tract. *Task names:* reading comprehension, sentence completion, verb generation, finger tapping, lip puckering.

| Patients |  |  |  |  | Fascicles in edema |  |  |  |  |  | fMRI tasks |  |  |  |  |
| --- | --- | --- | --- | --- | --- | --- | --- | --- | --- | --- | --- | --- | --- | --- | --- |
| Age range | Sex | Tumor type | Grade | L/R | AF | SLF3 | IFOF | FAT | CST | CBT | reading | sentence | verb | finger | lip |
| 36-40 | M | Astrocytoma | 2 | L | • | • |  | • | • | • | • | • | • | • | • |
| 66-70 | M | Astrocytoma | undef. | R | • | • | • | • | • | • |  |  |  | • |  |
| 51-55 | M | Glioblastoma | 4 | L | • | • |  | • | • | • | • | • | • |  | • |
| 66-70 | F | Glioblastoma | 4 | R | • | • | • | • | • |  |  | • | • | • | • |
| 41-45 | M | Astrocytoma | 3 | L | • | • | • | • | • | • | • | • | • | • | • |
| 41-45 | M | Oligodendroglioma | 3 | L | • | • |  | • | • | • | • | • | • | • | • |
| 71-75 | F | Oligodendroglioma | undef. | L | • | • | • | • | • | • |  | • | • | • | • |
| 56-60 | M | Glioblastoma | 4 | R | • | • | • |  | • |  | • | • | • |  |  |
| 31-35 | M | Oligodendroglioma | 2 | R | • | • |  |  |  |  | • | • | • | • |  |
| 71-75 | M | Oligodendroglioma | 3 | L | • | • | • | • | • | • | • | • | • | • | • |
| 61-65 | M | Oligodendroglioma | 3 | R | • | • | • | • | • |  |  |  |  | • |  |
| 51-55 | M | Glioblastoma | 4 | R | • | • | • | • |  | • |  | • | • | • |  |
| 41-45 | F | Glioblastoma | 4 | L | • | • | • | • | • | • | • | • | • | • | • |
| 51-55 | M | Oligodendroglioma | 3 | L | • | • | • | • |  |  | • | • | • | • | • |
| 41-45 | F | Astrocytoma | 3 | L | • | • |  | • | • |  | • | • | • | • | • |
| 46-50 | M | Glioblastoma | 4 | L | • | • | • | • | • |  | • | • | • | • | • |
| 51-55 | M | Glioblastoma | 4 | R | • | • |  | • | • | • | • | • | • | • | • |
| 31-35 | F | Oligodendroglioma | 3 | R | • | • | • | • | • | • | • | • | • | • |  |
| 61-65 | F | Glioblastoma | 4 | L | • | • | • |  |  |  | • | • | • |  | • |

#### 3.1. Diffusion MRI

##### Acquisition

The diffusion-weighted images (DWIs) were acquired with either 1.7 × 1.7 × 3.0 mm^3^ or 2.0 × 2.0 × 2.0 mm^3^ voxel size; echo time TE = 92 ms; repetition time TR = 5900 ms; 3 b-shells with (20,60,60) diffusion-encoding directions sampled, respectively, at *b* = 1000, 2500, 5000 s/mm^2^, interleaved with 6 images at *b* = 0, and followed by another *b* = 0 image acquired with the opposite (posterior→anterior) phase encoding.

##### Data preprocessing

Our data processing pipeline in MRtrix3 [28] consisted of Marchenko-Pastur PCA denoising (`dwidenoise`) [29] followed by removal of Gibbs ringing (`mrdegibbs`) [30], B1 field inhomogeneity (`dwibiascorrect ants`) [31], motion and eddy current artifacts (`dwifslpreproc`) [32]. After these steps, we resliced the images (`mrgrid` interpolation with Gaussian smoothing) to ensure consistent isotropic 2 × 2 × 2 mm^3^ voxel size across all DWIs. Finally, we manually drew the edema regions covering the areas of peritumoral hyperintensity in FLAIR images registered to DWIs with affine transitions.

##### ODF fingerprinting

We executed ODF-FP with a randomly sampled [6] dictionary of 10^6^ elements (i.e., one million) having 0 ≤ *N_j_* ≤ 3 fibers per voxel, generated within the default ranges of microstructure parameters (Table 2).

**Table 2:** Microstructure parameters used for generating ODF-dictionaries.

| Fraction sizes |  | Range |
| --- | --- | --- |
| $p_{\text{fib}}$ | fiber fraction | [0, 1] |
| $p_{\text{iso}}$ | isotropic fraction | [0, 1] |
| $f_{\text{in}}$ | intra-axonal fraction | [0, 1] |
| Diffusivities |  | Range [m<sup>2</sup>/s] |
| $D_a$ | intra-axonal | $[1.5, 2.5] \cdot 10^{-9}$ |
| $D_e^{\parallel}$ | extra-axonal parallel | $[1.5, 2.5] \cdot 10^{-9}$ |
| $D_e^{\perp}$ | extra-axonal perpendicular | $[0.5, 1.5] \cdot 10^{-9}$ |
| $D_{\text{iso}}$ | isotropic | $[2.0, 3.0] \cdot 10^{-9}$ |

For ODF-FP matching, we empirically chose the spurious fibers penalty factor λ = 2 · 10^−5^ and the anisotropy boosting factor µ = 0.1 throughout the whole brain. On top of that, we tested an exploratory set of anisotropy boosting factors µ_edema_ ∈ {0.1, 0.15, 0.2, 0.25, 0.3, 0.35, 0.4, 0.45, 0.5}used only inside the manually drawn regions of edema. For simplicity of notation, we will refer to these cases as ODF-FP_0.1_, ODF-FP_0.15_, …, ODF-FP_0.5_. Finally, we ran ODF-FP without any boosting either inside or outside edema (ODF-FP_no boost_).

The Python source code of ODF-FP is available at:https://github.com/filipp02/odffp *Standard methods.* For comparison, we processed the same DWIs using GQI, Functional magnetic resonance imaging of the brain Software Library (FSL) Bedpostx [33], and CSD with the Multi-Shell Multi-Tissue option (CSD-MSMT) [34].

##### Clinically-feasible methods

To assess the potential for clinical translation, we tested the tractography performance on subsampled single-shell data. We discarded the two highest b-shells (i.e., 2500 and 5000 s/mm^2^), thus restricting the imaging protocol to 20 DWIs at b=1000 s/mm^2^, which contemporary clinical scanners can acquire within a few minutes. We then executed:

- ODF-FP, GQI, and FSL with no changes compared to the fully sampled data set;
- CSD without the MSMT option (due to the missing higher b-shells);
- FWE-DTI and FERNET in their original implementations dedicated for single-shell data.

##### Tractography

We applied the Euler’s-method-based deterministic tractography algorithm [35] implemented in DSI Studio [36]. As the stopping criterion, we set the threshold on Normalized Quantitative Anisotropy (NQA) at 0.10. Note that NQA is the Quantitative Anisotropy (QA) [24, 36] measure scaled to the [0,1] range, which we used to ensure consistency among all tested subjects. Additionally, we limited the number of random seeds to 1 million, we set the maximum turning angle of 60^°^, and restricted the streamline lengths to the range between 30 and 200 mm. To dissect the fascicles traversing edema (Table 1), we used the atlas-based tracking in DSI Studio [37] with the tolerance of 30 mm aimed to accommodate the variability of tissue impacted by tumor.

### 3.2. Functional MRI

#### Acquisition

The task-based fMRI was acquired with 2.3 × 2.3 × 2.3 mm^3^ voxel size; echo time TE = 30 ms; and repetition time TR = 2000 ms. The tasks were chosen based on the tumor location and were grouped into the following categories (Table 1):

- **language tasks** — reading comprehension (14 patients), sentence completion (17), and verb generation (17);
- **motor tasks** — finger tapping contralateral to the tumor location (16 patients) and lip puckering (13).

The patients were trained before imaging, then asked to repeat each task 4 times in alternating 20-second periods of task and rest. Each sequence took 160 seconds, i.e., 4 × (20s task + 20s rest).

#### Data preprocessing

We preprocessed the data using fMRIprep [38] with default settings. Next, we completed the first-level analysis in fMRI Expert Analysis Tool (FEAT) [39] with Z-threshold=3.5 to obtain Blood Oxygenation Level Dependent (BOLD) maps. The tracking algorithm was not informed about BOLD activation regions.

### 3.3. Evaluation

For each WM fiber reconstruction method *m* (executions of the same algorithm with fully sampled and subsampled data sets were treated as separate methods), we dissected the fascicles traversing edema (Table 1).

To quantify each method’s ability to track through edematous WM, we calculated normalized edema overlap enhancement (OE) defined as:

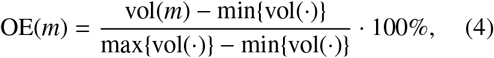

where

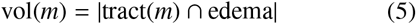

is the volume of the overlap between a tract dissected with the method *m* and the peritumoral vasogenic edema region drawn manually by a trained expert.

Note that min {vol(·)} and max{vol(·)} are, respec-tively, the lowest and the highest volumes among all tested methods per subject. Thus, for each patient and for each tract, OE(*m*) varied between 0% (worst-performing method) and 100% (best-performing method).

Analogously, we quantified the fMRI reference agreement using BOLD maps in the following metrics:

- **True Positive Ratio** (TPR):

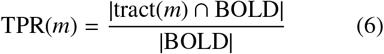
- **Dice Coe**ffi**cient** (DC):

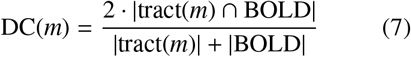
- **95% Hausdor**ff **distance** (HD_95_):

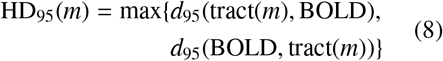

Where

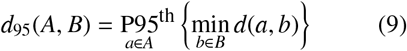

is the 95^th^ percentile of the distances between the elements of the sets *A* and *B*, i.e., either tract(*m*) or BOLD.

To compensate for the inter-subject variability, we normalized each of these metrics, as in Equation 4:

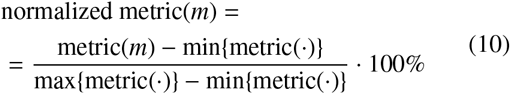

When computing the overlaps between tractography and BOLD maps, we coupled AF, SLF3, and FAT with all the language tasks, CST with the finger tapping task, and CBT with the lip puckering task.

Finally, we compared ODF-FP with the reference methods using the analysis of covariance (ANCOVA) at the significance level α = 0.05. As covariates, we considered: age, sex, tumor type and hemisphere.

## 4. Results

Our experiments showed that anisotropy boosting in ODF-FP had a visible impact on tractography (Figure 3). For all studied fascicles, the linear increment of µ_edema_ ≥ 0.1 gradually increased the edema OE, which began to stabilize within the 0.3–0.5 range. Figure 3 illustrates this pattern in the reconstructions of AF (Figure 3, upper panel) and the pyramidal tract, i.e., the CST/CBT complex (Figure 3, lower panel) immersed in peritumoral vasogenic edema.

**Figure 3.**
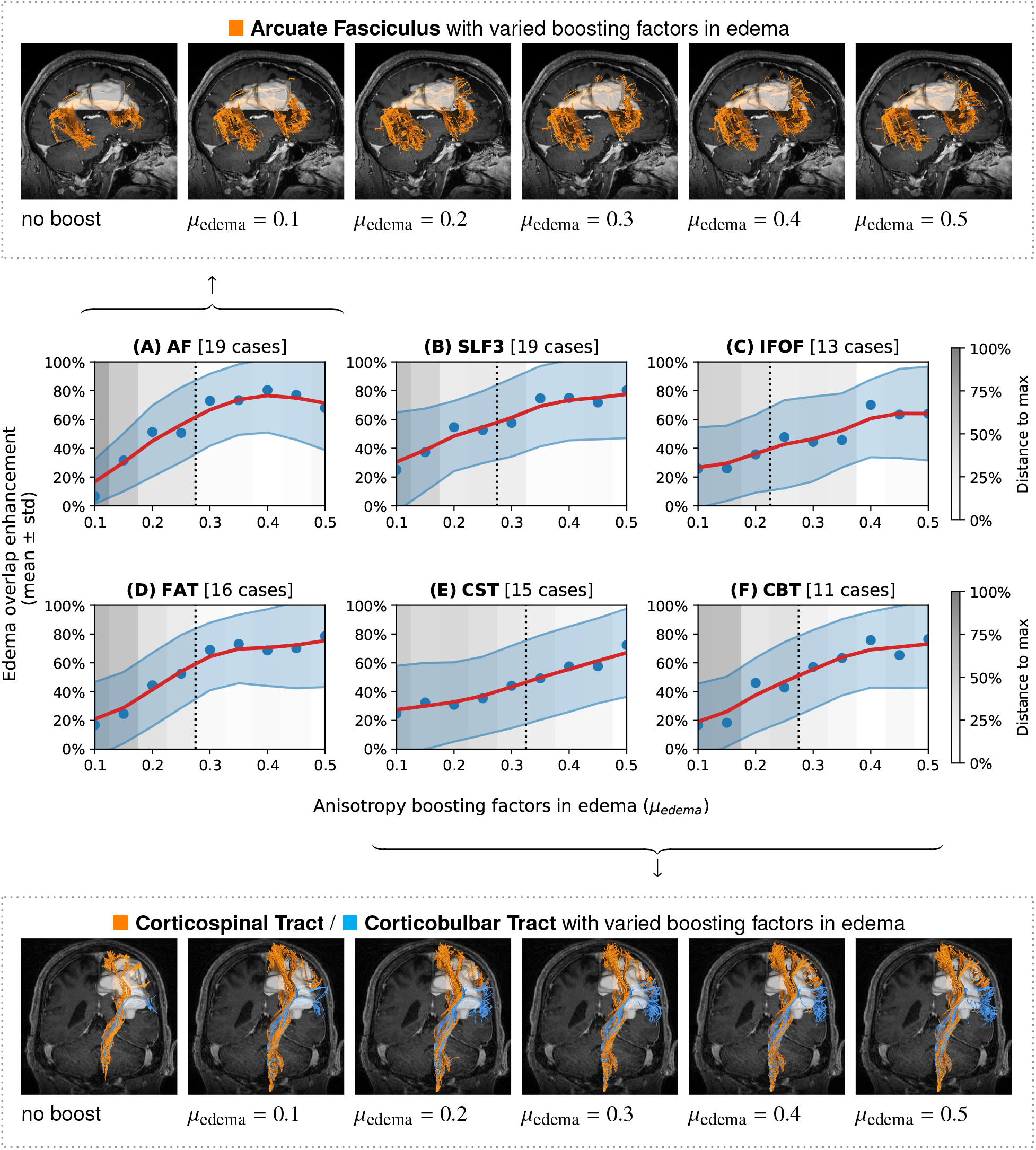
The impact of varied anisotropy boosting factors in edema (0.1 ≤ µedema ≤ 0.5) on tracking with ODF-FP. The plots at the center show relative volume overlaps between tractography and edema regions (blue dots) with regressed averages (red curves) and standard deviations (light blue areas) to illustrate the trends. The gray scale shows relative distances to the best-performing variant of µedema. The overlaps stabilized within 25% from the maximum (dotted line) or the approximate 0.3 ≤ µedema ≤ 0.5 range. Detailed views are presented for AF (top row) and CST/CBT (bottom row). Abbreviations: AF – Arcuate Fasciculus, SLF3 – Superior Longitudinal Fasciculus III, IFOF — Inferior Fronto-Occipital Fasciculus, FAT — Frontal Aslant Tract, CST — Corticospinal Tract, CBT — Corticobulbar Tract.

### 4.1. Overlap between tractography and edema

The mean overlap between tractography and edema was significantly higher in ODF-FP_0.5_ than in the standard methods (Figure 4), both in the fully sampled (*p* ≤ 0.03) and the subsampled data sets (*p* ≤ 0.05). For instance, our reconstruction of AF traversing edema in 19 patients with the fully sampled dMRI reached OE(*ODF-FP*_0.5_) = 74 ±28% (mean ± std), whereas the best-performing reference method, FSL, scored OE(*FSL*) = 45 ± 22%. In other words, on average, 74% of the edematous WM volume pertaining to AF — tracked with the combined effort of all tested methods — was *de facto* reconstructed with ODF-FP_0.5_, while FSL reconstructed 45% of this volume.

**Figure 4.**
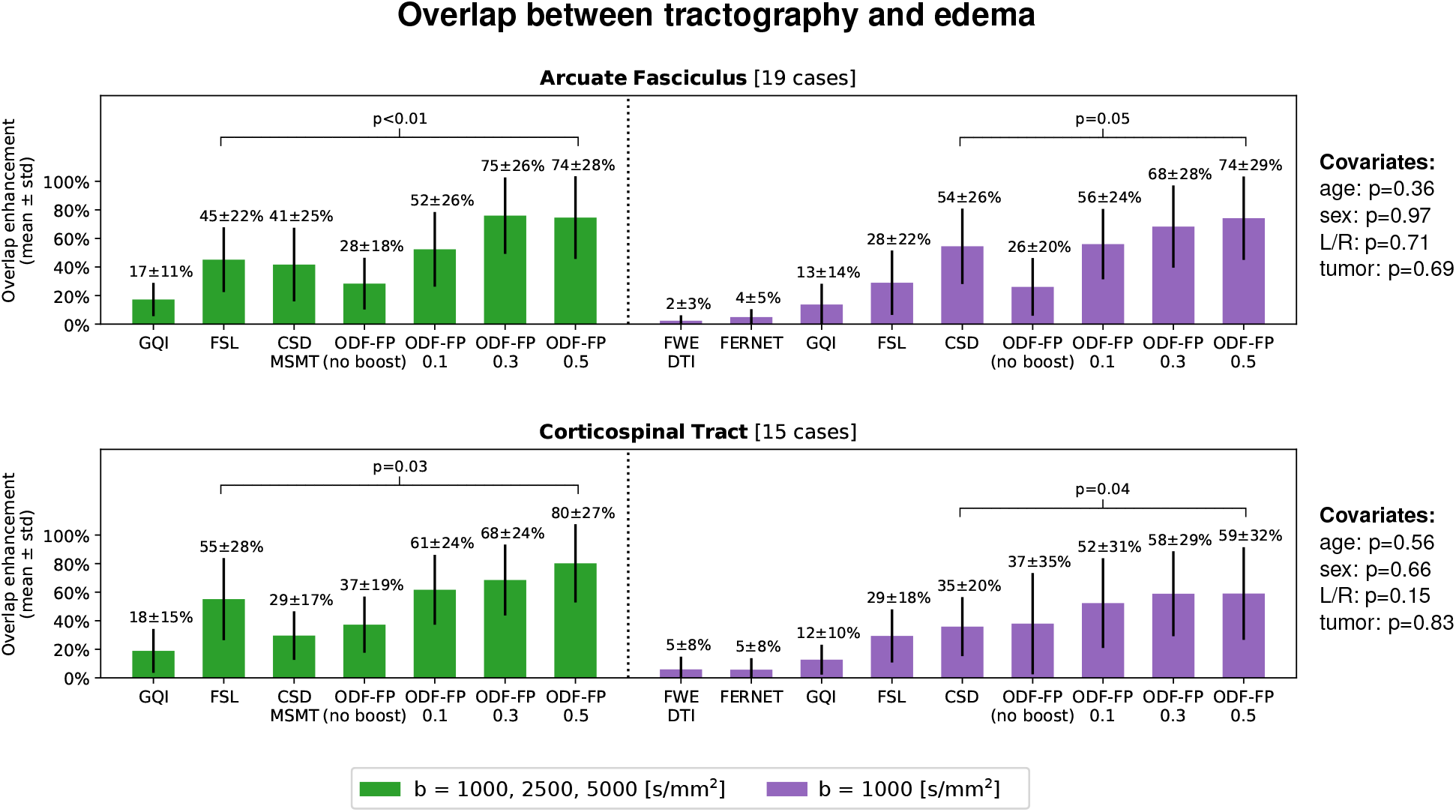
The relative overlap between tractography and edema (mean and standard deviation) in the fully sampled (green) and the subsampled clinically-feasible data sets (purple) calculated for Arcuate Fasciculus and Corticospinal Tract. ODF-FP is compared with the best-performing reference method, i.e., FSL in the fully sampled and CSD in the subsampled data set. The p-values of the covariates (patient’s age and sex, tumor hemisphere and tumor type) are given on the right side.

For the single-shell data, ODF-FP_0.5_ maintained an almost identical overlap between AF and edema (74 ± 29%). The best-performing reference method in the subsampled data set was CSD (56 ± 24%), while the tensor-based methods produced the lowest OEs: 2 ± 3% (FWE-DTI) and 4 ± 5% (FERNET).

It is also worth noticing that the impact of covariates on the overlap between tractography and edema was negligible, with the lowest p-value of 0.36 attributed to the age of patients.

Similar patterns in the overlap measures and the low impact of covariates were observed in other fascicles traversing peritumoral vasogenic edema: CST (Figure 4), SLF3, and FAT (Figure S1 in the Supplementary material). Exceptions were IFOF (13 cases) and CBT (11 cases), where ODF-FP_0.5_ still outperformed the standard methods, although the differences in OE were not statistically significant (Figure S1), which may relate to the relatively lower numbers of cases.

Figure 5 shows a representative example of the pyramidal tract traversing a large region of vasogenic edema. While all tested methods visualized the cortical projections of the tract located near the midline, only CSD, FSL, and ODF-FP reconstructed the lateral branches most impacted by edema. Among these branches, our method visualized the biggest part of the CBT.

**Figure 5.**
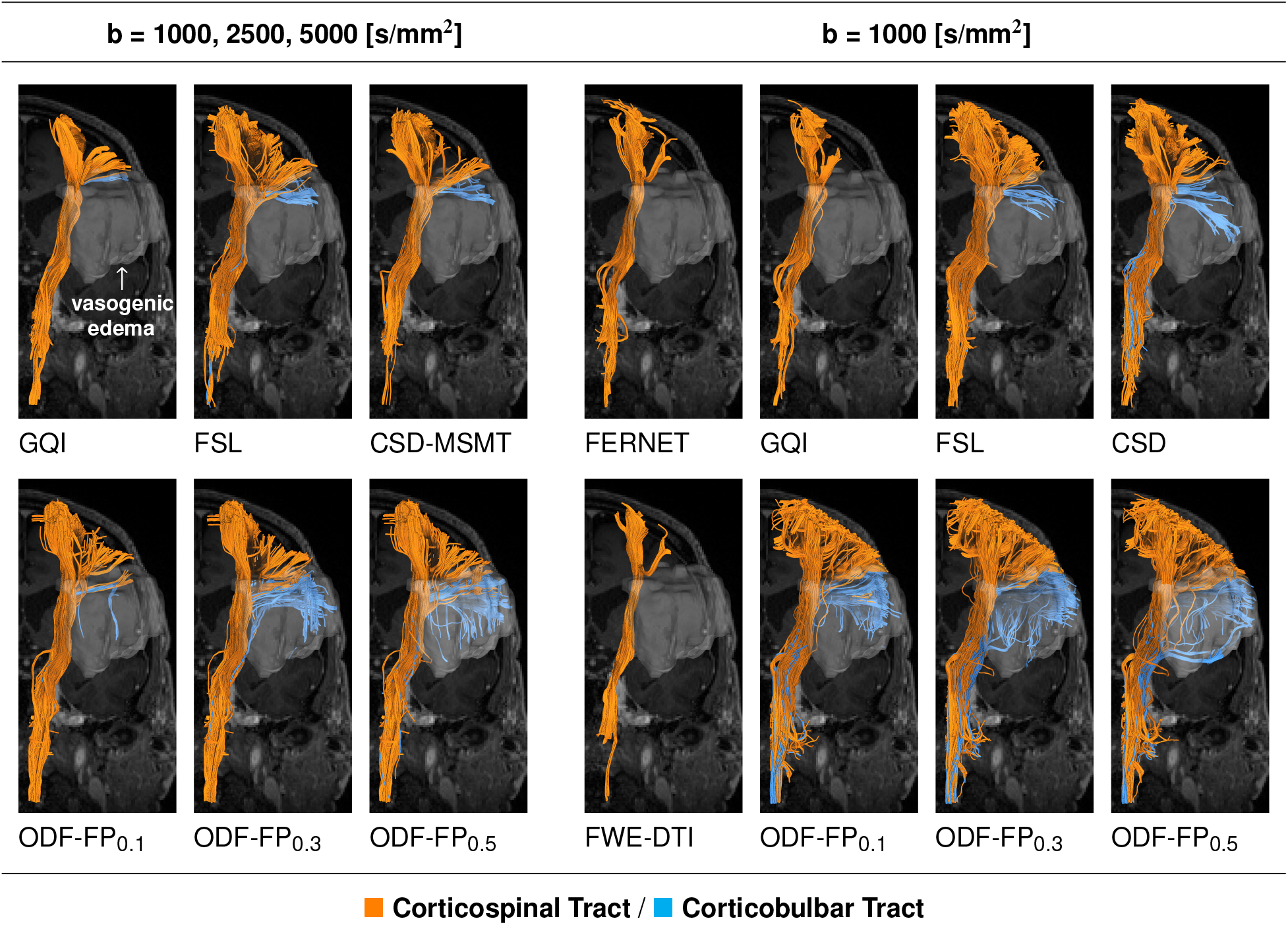
Tractography reconstruction of the pyramidal tract — composed of Corticospinal (CST, orange streamlines) and Corticobulbar Tract (CBT, blue streamlines) — traversing a significant vasogenic edema. All tested methods visualized the cortical projections of the pyramidal tract located near the midline. ODF-FP reconstructed the highest volume of CBT immersed in edema, both in the fully sampled (left group of columns) and the subsampled dMRI (right group of columns).

Also, note that data subsampling did not cause ODFFP to produce less streamlines. However, it resulted in a slightly more winding shape of the reconstructed tracts and an occurrence of potential false positives at µ_edema_ = 0.5 (Figure 5).

### 4.2. Overlap between tractography and fMRI

Measuring the overlap between CST tractography and BOLD maps computed from the finger tapping task did not reveal a single best-performing method in the fully sampled data set. However, the superiority of ODF-FP over the standard methods was significant (*p* < 0.01) in the single-shell dMRI (Figure 6).

**Figure 6.**
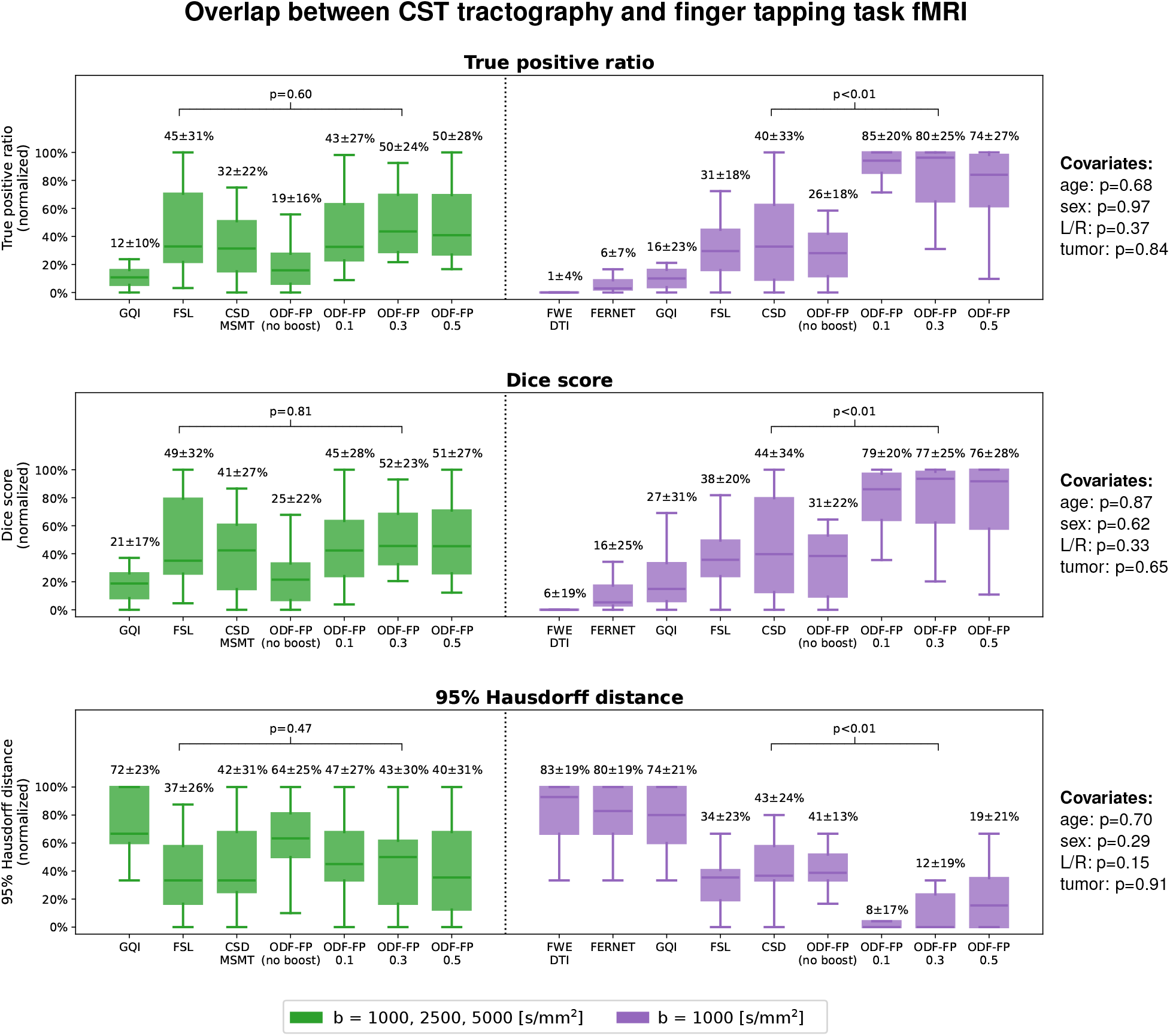
Box plots of the relative overlap between Corticospinal Tract (CST) and the cortical area activated during the finger tapping functional MRI (fMRI) task. Tractography was produced from the fully sampled (green) or the subsampled clinically-feasible diffusion MRI (purple). The mean and standard deviations of the normalized True positive rates, Dice scores, and 95% Hausdorff distances are given above the respective box whiskers. ODF-FP is compared with the best-performing reference method, i.e., FSL in the fully sampled and CSD in the subsampled data set. The p-values of the covariates (patient’s age and sex, tumor hemisphere and tumor type) are given on the right side.

Unlike the overlap with edema, the agreement between tractography and fMRI did not relate proportionally to the anisotropy boosting. Particularly, ODF-FP_0.3_ outperformed ODF-FP_0.5_, i.e., reached higher TPR, higher DC, and lower HD95 (Figures 6 and S2), which reinforced the earlier hypothesis of false positives at µ_edema_ = 0.5.

Figure 7 shows a coronal slice through the peritumoral zone with the BOLD activation in the motor cortex acquired during the finger tapping task. In this representative example, the CST reconstructions based on GQI, FERNET, FWE-DTI, and CSD-MSMT terminated tracking before reaching most of the cortical terminations in the region highlighted with fMRI, whereas FSL, CSD, and ODF-FP reached a visible overlap with the BOLD activation region. Among them, ODF-FP reconstructed the wide-spread fanning shape of the CST most densely, both in the fully sampled (ODF-FP_0.5_) and the subsampled data set (all ODF-FP variants).

**Figure 7.**
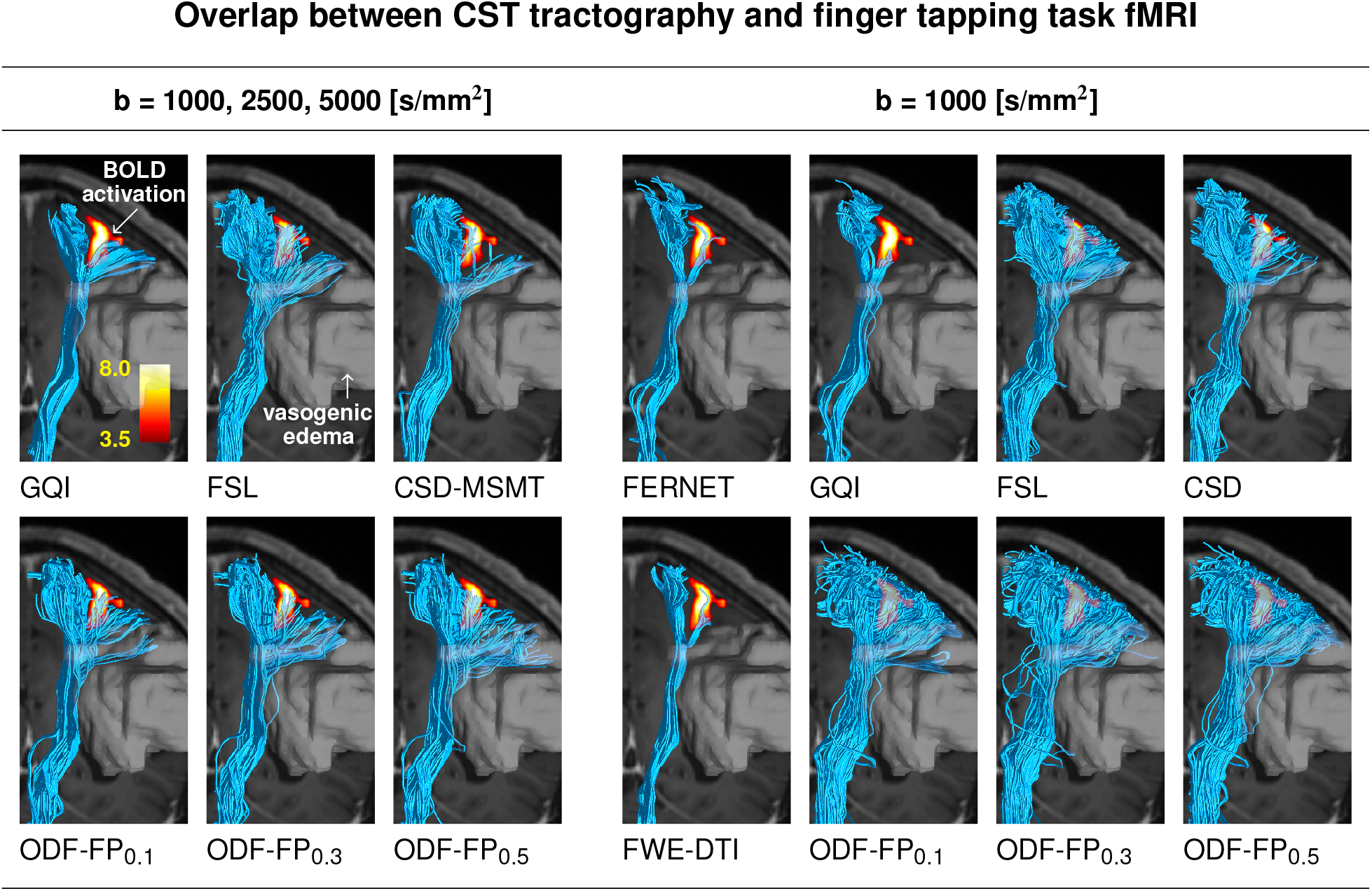
Coronal view of Corticospinal Tract (CST) overlapping with the cortical region activated during the finger tapping functional MRI (fMRI) task. Tractography was produced from the fully sampled (left group of columns) or the subsampled diffusion MRI (right group of columns). The tract traverses peritumoral vasogenic edema (white translucent region). The reconstructions based on GQI, FERNET, FWE-DTI, and CSD-MSMT terminated tracking before reaching most of the cortical terminations in the region highlighted with fMRI, whereas FSL, CSD, and ODF-FP reached a visible overlap with the BOLD activation region. Among them, ODF-FP reconstructed the wide-spread fanning shape of the CST most densely, both in the fully sampled (ODF-FP_0.5_) and the subsampled data set (all ODF-FP variants).

In the language tasks, such as verb generation (Figure 8), ODF-FP typically reached higher agreement with fMRI than the standard methods, especially in the subsampled data sets. Interestingly, the patients’ age had a relatively big impact on the BOLD overlap with AF (Figure 8A) and SLF3 (Figure 8B), while sex — on the overlap with FAT (Figure 8C). Additional information is given in Supplementary Figures S3–S11.

**Figure 8.**
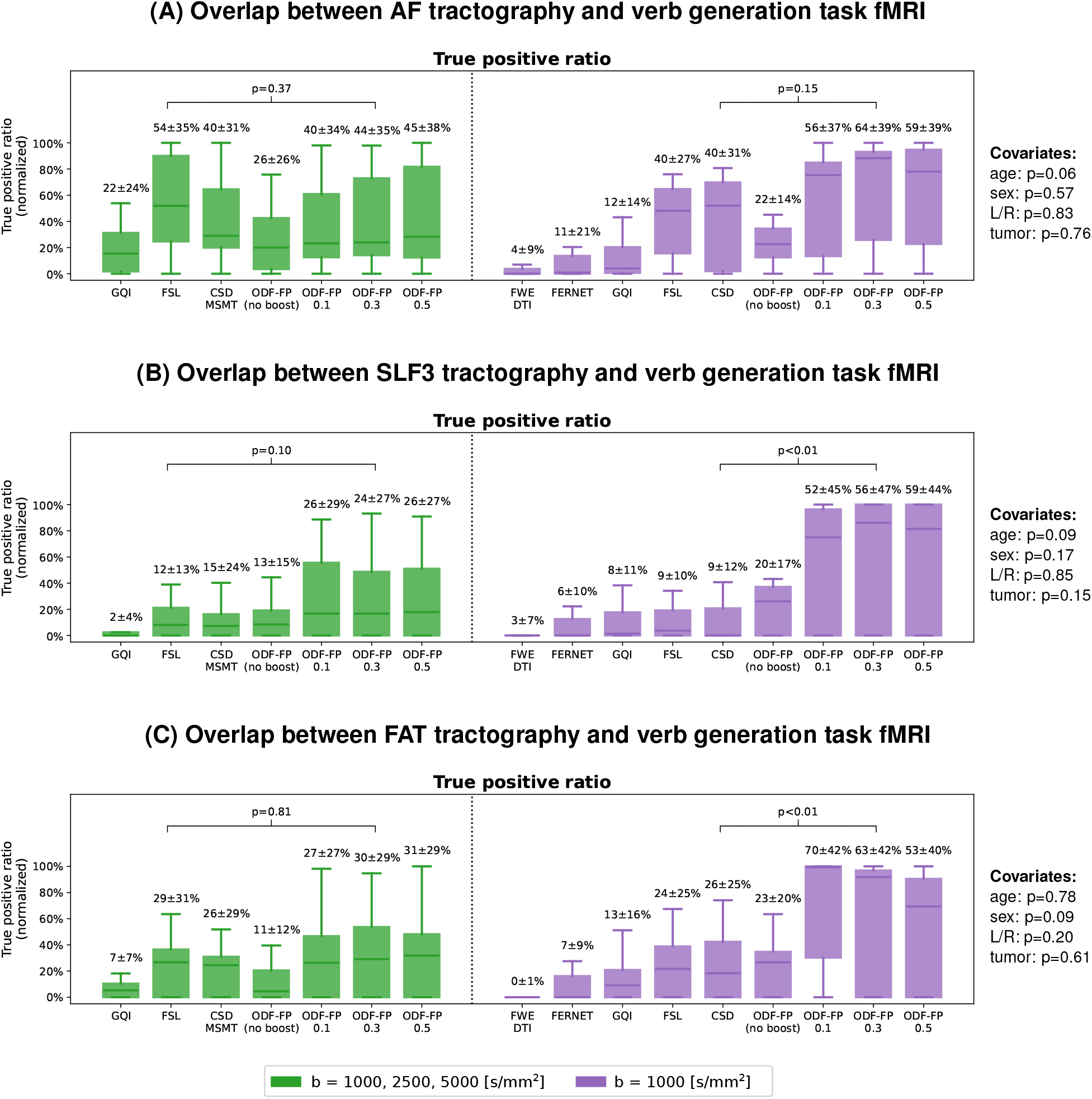
Box plots of the relative overlap between tractography of (A) AF – Arcuate Fasciculus, (B) SLF3 — Superior Longitudinal Fasciculus III, (C) FAT — Frontal Aslant Tract, and the cortical area activated during the verb generation functional MRI (fMRI) task. Tractography was produced from the fully sampled (green) or the subsampled clinically-feasible diffusion MRI (purple). The mean and standard deviations of the normalized True positive rates are given above the respective box whiskers. ODF-FP is compared with the best-performing reference method, i.e., FSL in the fully sampled and CSD in the subsampled data set. The p-values of the covariates (patient’s age and sex, tumor hemisphere and tumor type) are given on the right side.

Finally, it is worth noticing that maximizing the agreement between tractography and fMRI was more challenging in the case of AF (Figure 9), which overlapped with the corresponding BOLD maps in two distant regions (i.e., the anterior and posterior language areas), while all other combinations of tracts and BOLD maps overlapped in a single brain region. The AF reconstruction illustrated in Figure 9 was completed success-fully by FSL and ODF-FP in the fully sampled data set, although it posed a challenge in the subsampled data set, where only ODF-FP reconstructed the part of the tract located in the temporal lobe.

**Figure 9.**
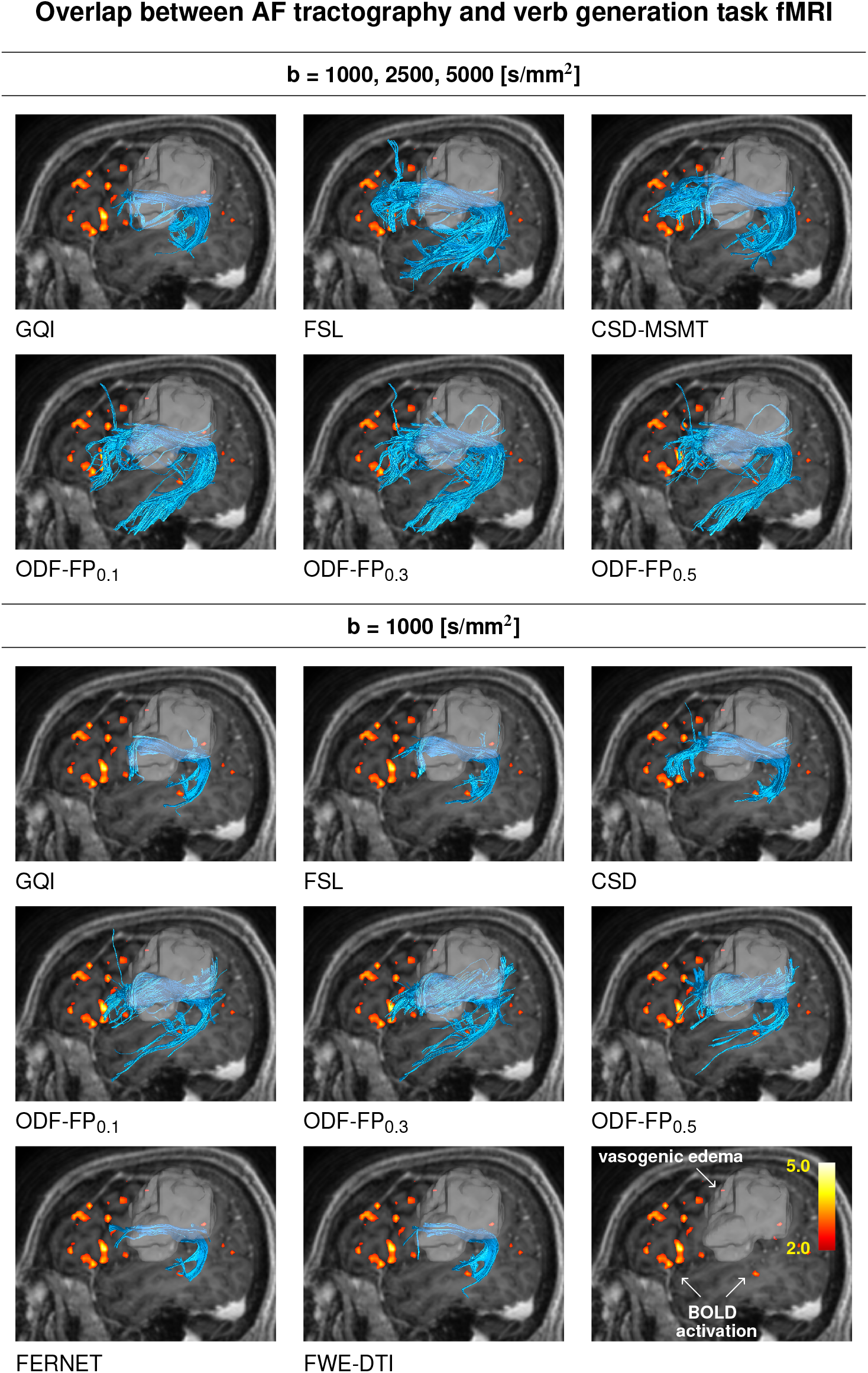
Sagittal view of Arcuate Fasciculus (AF) overlapping with the cortical region activated during the verb generation functional MRI (fMRI) task. Tractography was produced from the fully sampled (upper panel) or the subsampled diffusion MRI (lower panel). The tract traverses peritumoral vasogenic edema (white translucent region). The reconstruction was completed successfully by FSL and ODF-FP in the fully sampled data set, although it posed a challenge in the subsampled data set, where only ODF-FP reconstructed the part of the tract located in the temporal lobe.

## 5. Discussion

The accuracy of tractography varies throughout the brain parenchyma [40] with significant errors observed in peritumoral vasogenic edema [1]. Our experiments show that ODF-FP with the proposed diffusion anisotropy boosting mechanism can be robust to signal distortions in edematous WM.

### 5.1. Clinical utility

The standard of care in glioma treatment requires maximal safe resection [12, 41]. However, neurosurgeons lack accurate structural imaging modalities to support their decision regarding the resection volume when vasogenic peritumoral edema distorts WM tractography.

In this paper, we demonstrate that ODF-FP can outperform the standard fiber reconstruction methods when executed on research-quality densely sampled DWIs as well as the clinically-feasible single-shell data with 20 encoding directions at *b* = 1000 s/mm^2^. Especially the latter aspect carries the potential for clinical translation. Upon further validation, our approach could improve reconstruction of WM fiber tracts in various conditions beyond brain tumors, such as stroke or traumatic brain injuries, where edema limits diagnostic reliability of tractography.

### 5.2. Practical recommendations

As with many regularization parameters, choosing the anisotropy boosting factor may require a trial and error procedure. In our experiments, 0.3 ≤ µ_edema_ ≤ 0.5 proved most efficient. Particularly, µ_edema_ = 0.5 maximized the overlap between tractography and edema, while µ_edema_ = 0.3 the overlap with fMRI. The risk of false positives additionally inclines toward choosing the lower values (such as µ_edema_ = 0.3), although the optimal selection requires adjustment to each data set.

It is also worth noticing that the variant with the same anisotropy boosting factor used in the whole brain, i.e. either inside or outside edema (µ = µ_edema_ = 0.1), ensured relatively high performance. In practice, such a variant may help to streamline data processing, as it does not require drawing an edema ROI.

### 5.3. Measurement limitations

Quantification of clinical tractography is inherently limited due to the lack of ground truth. Therefore, all evaluation measures presented in this paper are indirect. The *edema OE* metric, introduced here, is an analog of the *bundle fraction in edema* used by Deslauriers-Gauthier et al. [42]. Both these metrics are based on the elementary assumption that improvement in tracking across edematous WM increases the overlap between tractography and edema. Hence, intuitively: the more robust tractography, the higher OE. However, the opposite relation (i.e., that higher overlap implies improvement in tractography) may not hold due to false positives. In the extreme scenario, the edema OE metric by definition reaches its maximum for a combination of all tracking methods being compared, although such a solution is suboptimal, because it also maximizes the number of false positives.

To address this issue, we included BOLD maps of the language-or motor-related tasks. By matching them with the respective WM tracts involved in the language (AF, SLF3, IFOF) or motor functions (CST, CBT), we identified the cortical areas that these tracts were structurally connected to. Consequently, the TPR of the overlap between tractography and BOLD maps helped us estimate the accuracy of tracking toward the activated cortical areas (since higher overlap implied that more distinct streamlines reached the target). Analogously, DC and HD95 implicitly estimated the impact of potential false positives, assuming that spurious fibers would deviate or drift away from the actual tract, therefore decreasing DC and increasing HD95. However, our BOLD maps showed activation in all regions involved in the performed tasks (such as visual cortex or premotor cortex), not all of which were structurally connected to the reconstructed tracts. From this perspective, fMRI-based validation is biased by potential false positives.

Additional limitations arose from the relatively low number of patients (with respect to the variability of glioma) and inconsistent voxel sizes in the acquired dMRI. Also, note that the use atlas-based tracking approach, which helped us improve consistency of the results, might have simultaneously limited the sensitivity to inter-subject variability.

### 5.4. Future work

The proposed approach is promising, although its clinical applicability requires further validation. Future work should employ a dedicated synthetic model of edematous tissue or a more precise reference measurement in vivo, such as intraoperative electrical stimulation of the brain, to ensure direct evaluation of the tracking accuracy.

## 6. Conclusion

We proposed a WM fiber reconstruction method to improve tracking in peritumoral zones with vasogenic edema. Our approach based on ODF-FP with a dedicated regularization term to boost diffusion anisotropy outperformed the standard methods by producing higher volumes of streamlines in the regions of vasogenic edema and by reaching higher overlaps with the cortical regions activated during task-based fMRI. The proposed method also proved effective when applied to clinically-feasible single-shell dMRI, which offers an opportunity for wide applicability in surgical planning and interventions.

## Data Availability

All data produced in the present study are available upon reasonable request to the authors.

## Acknowledgments

This project was supported in part by the National Institutes of Health (NIH, R01 EB028774 and R01 EB029306) under the rubric of the Center for Advanced Imaging Innovation and Research (CAI2R, https://www.cai2r.net), an NIBIB Biomedical Technology Resource Center (NIH P41 EB017183).

## Supplementary material

Supplementary Figure S1: The relative overlap between tractography and edema (mean and standard deviation) in the fully sampled (green) and the subsampled clinically-feasible data sets (purple) calculated for all the reconstructed fascicles. ODF-FP is compared with the best-performing reference method, i.e., FSL in the fully sampled and CSD in the subsampled data set. The p-values of the covariates (patient’s age and sex, tumor hemisphere and tumor type) are given on the right side.

Supplementary Figure S2: Box plots of the relative overlap between Corticobulbar Tract (CBT) and the cortical area activated during the lip puckering functional MRI (fMRI) task. Tractography was produced from the fully sampled (green) or the subsampled clinically-feasible diffusion MRI (purple). The mean and standard deviations of the normalized True positive rates, Dice scores, and 95% Hausdorff distances are given above the respective box whiskers. ODF-FP is compared with the best-performing reference method, i.e., FSL in the fully sampled and CSD in the subsampled data set. The p-values of the covariates (patient’s age and sex, tumor hemisphere and tumor type) are given on the right side.

Supplementary Figure S3: Box plots of the relative overlap between Arcuate Fasciculus (AF) and the cortical area activated during the reading functional MRI (fMRI) task. Tractography was produced from the fully sampled (green) or the subsampled clinically-feasible diffusion MRI (purple). The mean and standard deviations of the normalized True positive rates, Dice scores, and 95% Hausdorff distances are given above the respective box whiskers. ODF-FP is compared with the best-performing reference method, i.e., FSL in the fully sampled and CSD in the subsampled data set. The p-values of the covariates (patient’s age and sex, tumor hemisphere and tumor type) are given on the right side.

Supplementary Figure S4: Box plots of the relative overlap between Superior Longitudinal Fasciculus III (SLF3) and the cortical area activated during the reading functional MRI (fMRI) task. Tractography was produced from the fully sampled (green) or the subsampled clinically-feasible diffusion MRI (purple). The mean and standard deviations of the normalized True positive rates, Dice scores, and 95% Hausdorff distances are given above the respective box whiskers. ODF-FP is compared with the best-performing reference method, i.e., FSL in the fully sampled and CSD in the subsampled data set. The p-values of the covariates (patient’s age and sex, tumor hemisphere and tumor type) are given on the right side.

Supplementary Figure S5: Box plots of the relative overlap between Frontal Aslant Tract (FAT) and the cortical area activated during the reading functional MRI (fMRI) task. Tractography was produced from the fully sampled (green) or the subsampled clinically-feasible diffusion MRI (purple). The mean and standard deviations of the normalized True positive rates, Dice scores, and 95% Hausdorff distances are given above the respective box whiskers. ODF-FP is compared with the best-performing reference method, i.e., FSL in the fully sampled and CSD in the subsampled data set. The p-values of the covariates (patient’s age and sex, tumor hemisphere and tumor type) are given on the right side.

Supplementary Figure S6: Box plots of the relative overlap between Arcuate Fasciculus (AF) and the cortical area activated during the sentence completion functional MRI (fMRI) task. Tractography was produced from the fully sampled (green) or the subsampled clinically-feasible diffusion MRI (purple). The mean and standard deviations of the normalized True positive rates, Dice scores, and 95% Hausdorff distances are given above the respective box whiskers. ODF-FP is compared with the best-performing reference method, i.e., FSL in the fully sampled and CSD in the subsampled data set. The p-values of the covariates (patient’s age and sex, tumor hemisphere and tumor type) are given on the right side.

Supplementary Figure S7: Box plots of the relative overlap between Superior Longitudinal Fasciculus III (SLF3) and the cortical area activated during the sentence completion functional MRI (fMRI) task. Tractography was produced from the fully sampled (green) or the subsampled clinically-feasible diffusion MRI (purple). The mean and standard deviations of the normalized True positive rates, Dice scores, and 95% Hausdorff distances are given above the respective box whiskers. ODF-FP is compared with the best-performing reference method, i.e., FSL in the fully sampled and CSD in the subsampled data set. The p-values of the covariates (patient’s age and sex, tumor hemisphere and tumor type) are given on the right side.

Supplementary Figure S8: Box plots of the relative overlap between Frontal Aslant Tract (FAT) and the cortical area activated during the sentence completion functional MRI (fMRI) task. Tractography was produced from the fully sampled (green) or the subsampled clinically-feasible diffusion MRI (purple). The mean and standard deviations of the normalized True positive rates, Dice scores, and 95% Hausdorff distances are given above the respective box whiskers. ODF-FP is compared with the best-performing reference method, i.e., FSL in the fully sampled and CSD in the subsampled data set. The p-values of the covariates (patient’s age and sex, tumor hemisphere and tumor type) are given on the right side.

Supplementary Figure S9: Box plots of the relative overlap between Arcuate Fasciculus (AF) and the cortical area activated during the verb generation functional MRI (fMRI) task. Tractography was produced from the fully sampled (green) or the subsampled clinically-feasible diffusion MRI (purple). The mean and standard deviations of the normalized True positive rates, Dice scores, and 95% Hausdorff distances are given above the respective box whiskers. ODF-FP is compared with the best-performing reference method, i.e., FSL in the fully sampled and CSD in the subsampled data set. The p-values of the covariates (patient’s age and sex, tumor hemisphere and tumor type) are given on the right side.

Supplementary Figure S10: Box plots of the relative overlap between Superior Longitudinal Fasciculus III (SLF3) and the cortical area activated during the verb generation functional MRI (fMRI) task. Tractography was produced from the fully sampled (green) or the subsampled clinically-feasible diffusion MRI (purple). The mean and standard deviations of the normalized True positive rates, Dice scores, and 95% Hausdorff distances are given above the respective box whiskers. ODF-FP is compared with the best-performing reference method, i.e., FSL in the fully sampled and CSD in the subsampled data set. The p-values of the covariates (patient’s age and sex, tumor hemisphere and tumor type) are given on the right side.

Supplementary Figure S11: Box plots of the relative overlap between Frontal Aslant Tract (FAT) and the cortical area activated during the verb generation functional MRI (fMRI) task. Tractography was produced from the fully sampled (green) or the subsampled clinicallyfeasible diffusion MRI (purple). The mean and standard deviations of the normalized True positive rates, Dice scores, and 95% Hausdorff distances are given above the respective box whiskers. ODF-FP is compared with the best-performing reference method, i.e., FSL in the fully sampled and CSD in the subsampled data set. The p-values of the covariates (patient’s age and sex, tumor hemisphere and tumor type) are given on the right side.

## Notes

### Competing Interest Statement

The authors have declared no competing interest.

### Author Declarations

This retrospective study was approved by the Internal Review Board (IRB) of NYU Langone Health (protocol number i18-00124). Access to previously collected anonymized data did not require additional consent from the participants.

