## Supplementary material for "Clinically-feasible white matter fiber tractography in peritumoral zones with cerebral vasogenic edema"

Patryk Filipiak[a,*], Timothy M. Shepherd[a], Kamri Clarke[a], Dimitris G. Placantonakis[b], Fernando E. Boada[c], Steven H. Baete[a]

[a] *Center for Advanced Imaging Innovation and Research (CAI2R), Department of Radiology, NYU Langone Health, New York, NY, USA*
[b] *Department of Neurosurgery, Perlmutter Cancer Center, Neuroscience Institute, Kimmel Center for Stem Cell Biology, NYU Langone Health, New York, NY, USA*
[c] *Radiological Sciences Laboratory and Molecular Imaging Program at Stanford, Department of Radiology, Stanford University, Stanford, CA, USA*

**Abstract**

In diffusion MRI, vasogenic edema manifests as a major fraction of isotropic water that dilutes the anisotropic intra-axonal portion of the signal. Many tractography algorithms mistake vasogenic edema for the white matter boundary and terminate tracking to prevent producing spurious streamlines. As a result, visual representations of fascicles traversing edema are often compromised, limiting the clinical utility of tractography in neurosurgery.

We address this hurdle with ODF-Fingerprinting (ODF-FP) — a dictionary-based fiber reconstruction algorithm that accommodates variability of neural tissue. By adding a regularization term to the ODF-FP matching formula, we counterbalance the drop of diffusion anisotropy in edematous regions to improve white matter fiber identification.

We test our approach in 19 glioma cases with significant peritumoral vasogenic edema. For each case, we quantify the volume of the reconstructed white matter tracts immersed in edema, then we use the cortical regions activated during task-based functional MRI as validation for tractography. To assess the potential for clinical translation, we additionally test the performance of our reconstruction method on subsampled single-shell diffusion-weighted images, which contemporary clinical scanners can acquire within a few minutes.

*Corresponding author at: Center for Biomedical Imaging, NYU Langone Health, 660 1st Avenue, New York, NY 10016, USA

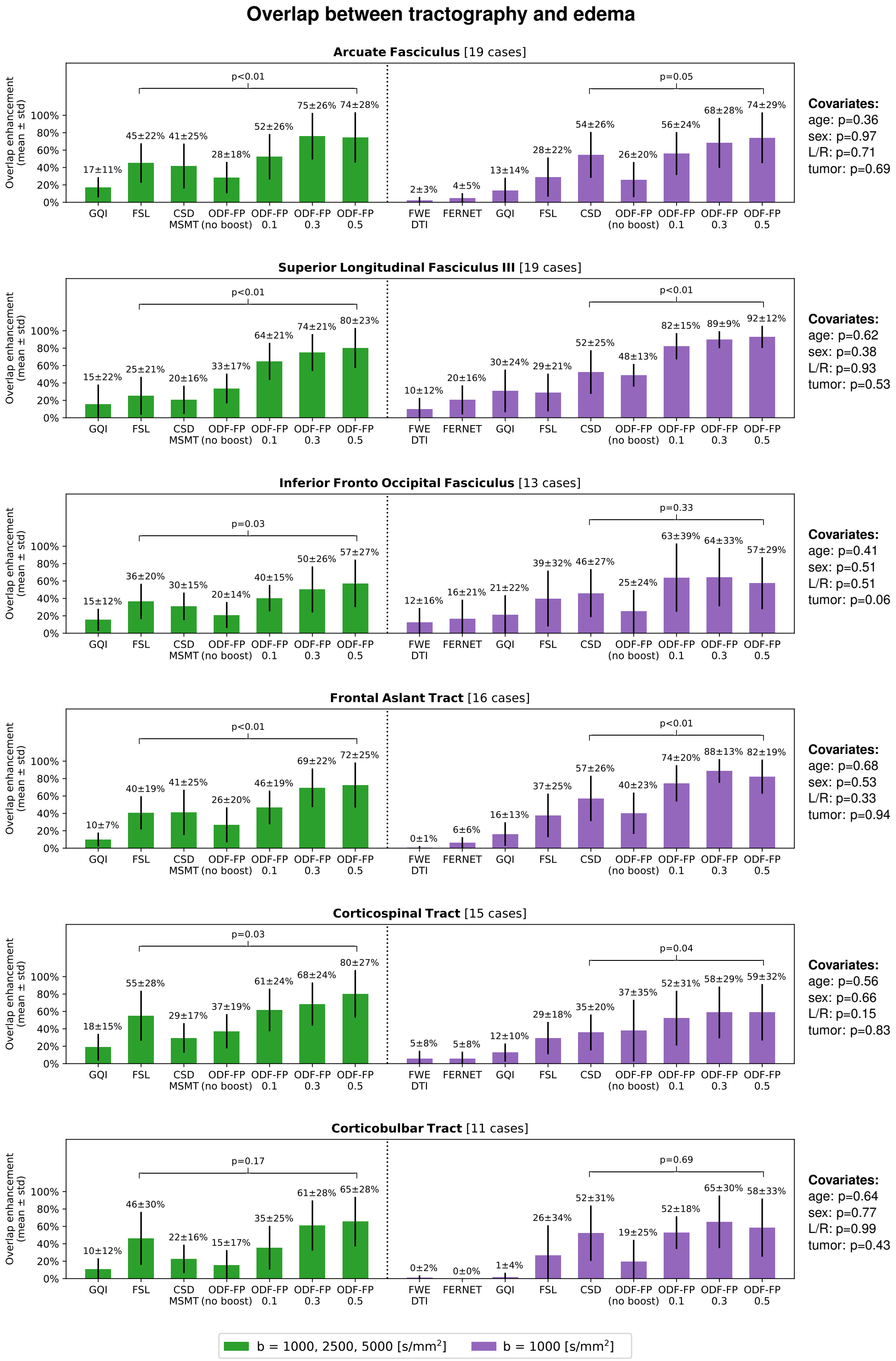


Supplementary Figure S1: The relative overlap between tractography and edema (mean and standard deviation) in the fully sampled (green) and the subsampled data sets (purple) calculated for all the reconstructed fascicles. ODF-FP is compared with the best-performing reference method, i.e., FSL in the fully sampled and CSD in the subsampled data set. The p-values of the covariates (patient's age and sex, tumor hemisphere and tumor type) are given on the right side.

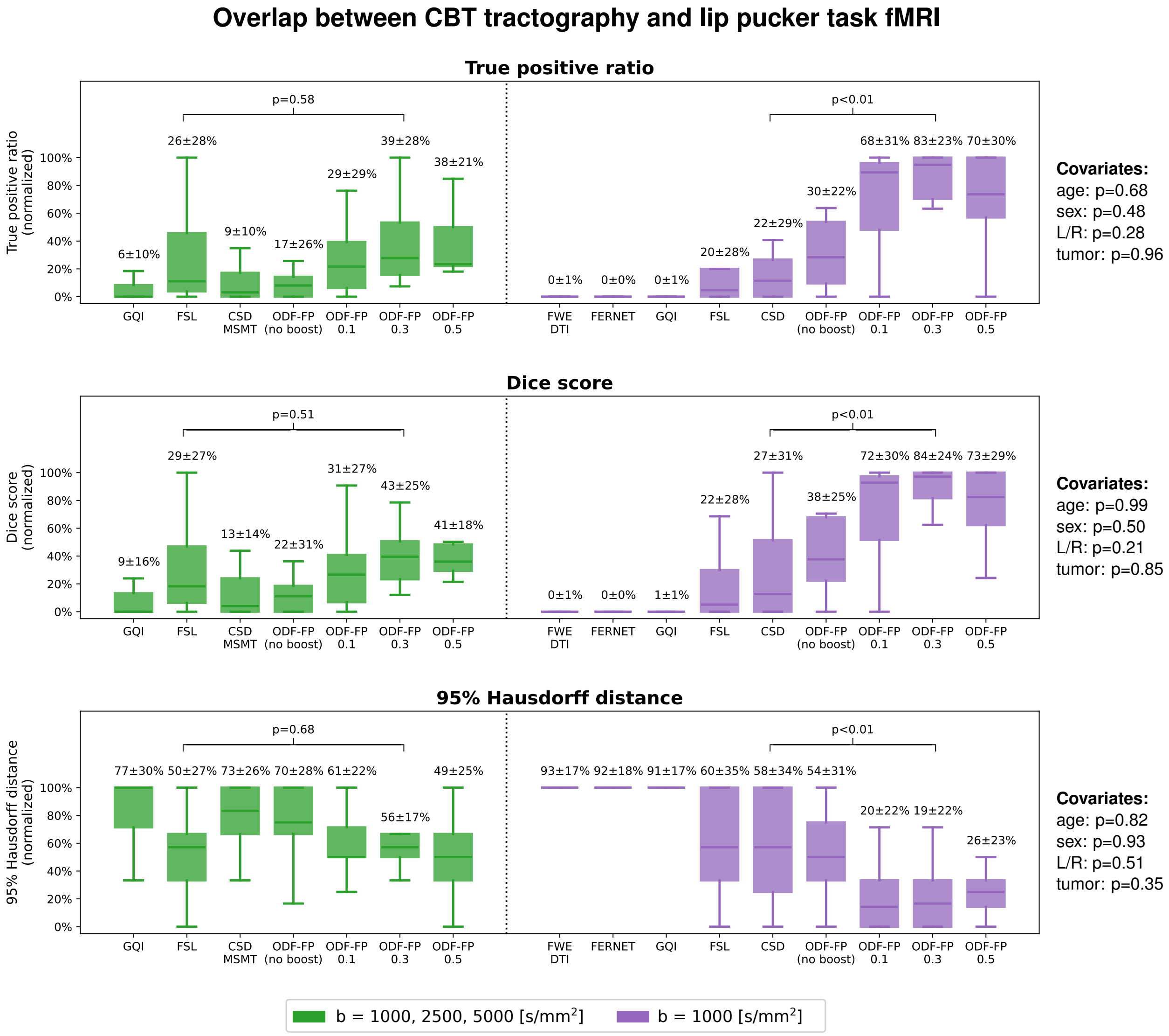


Supplementary Figure S2: Box plots of the relative overlap between Corticobulbar Tract (CBT) and the cortical area activated during the lip puckering functional MRI (fMRI) task. Tractography was produced from the fully sampled (green) or the subsampled clinically-feasible diffusion MRI (purple). The mean and standard deviations of the normalized True positive rates, Dice scores, and 95% Hausdorff distances are given above the respective box whiskers. ODF-FP is compared with the best-performing reference method, i.e., FSL in the fully sampled and CSD in the subsampled data set. The p-values of the covariates (patient's age and sex, tumor hemisphere and tumor type) are given on the right side.

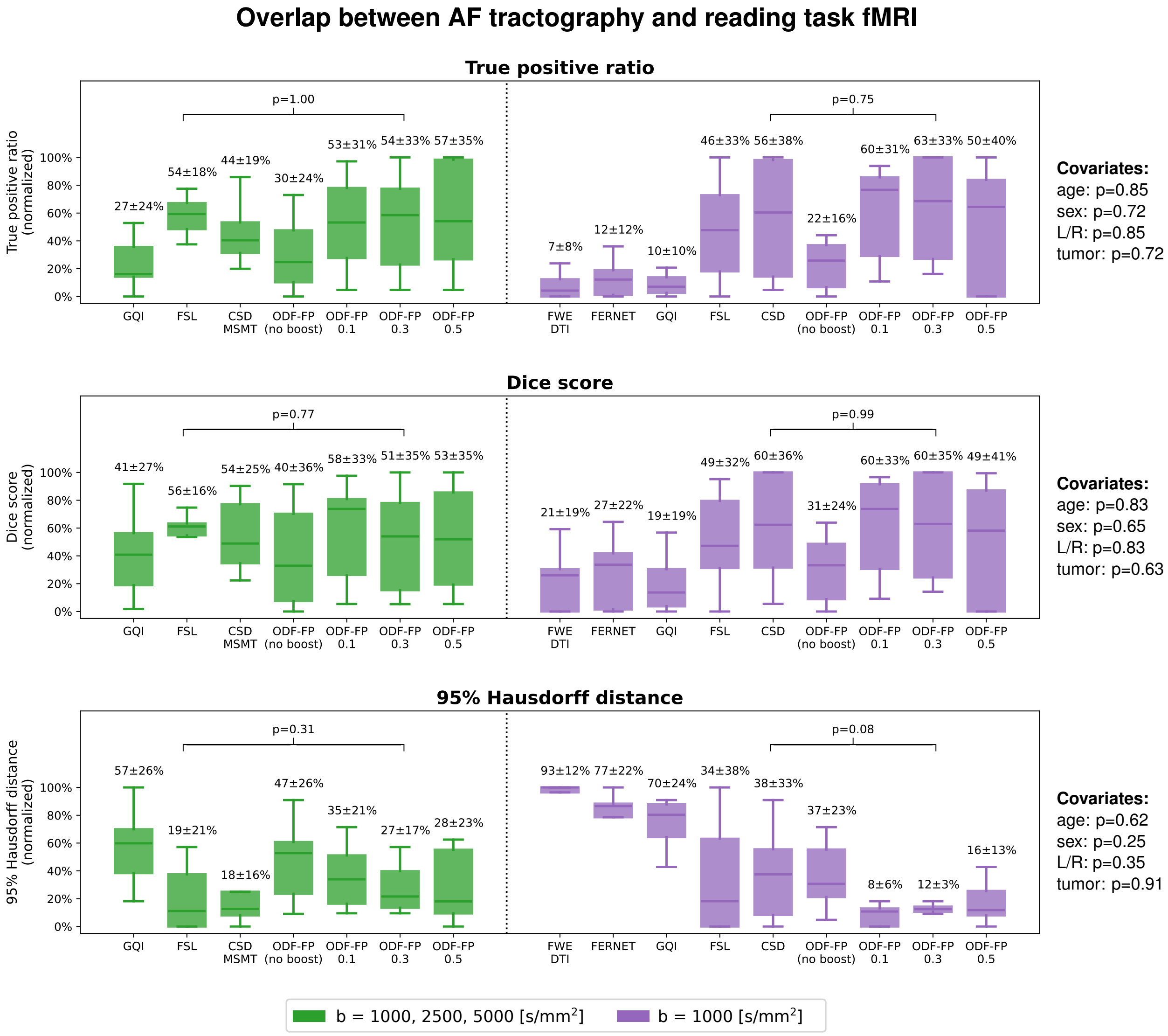


Supplementary Figure S3: Box plots of the relative overlap between Arcuate Fasciculus (AF) and the cortical area activated during the reading functional MRI (fMRI) task. Tractography was produced from the fully sampled (green) or the subsampled clinically-feasible diffusion MRI (purple). The mean and standard deviations of the normalized True positive rates, Dice scores, and 95% Hausdorff distances are given above the respective box whiskers. ODF-FP is compared with the best-performing reference method, i.e., FSL in the fully sampled and CSD in the subsampled data set. The p-values of the covariates (patient's age and sex, tumor hemisphere and tumor type) are given on the right side.

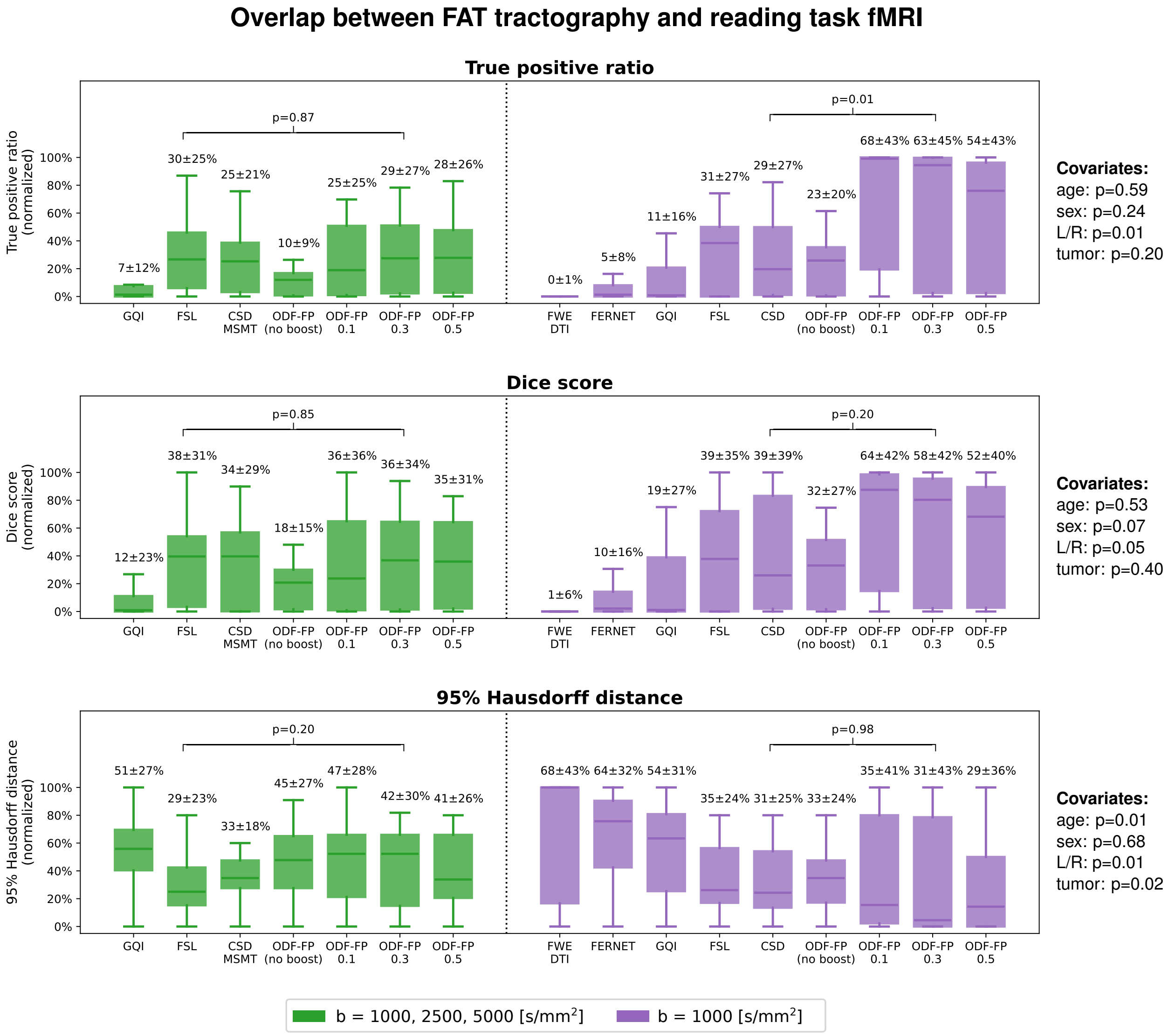


Supplementary Figure S4: Box plots of the relative overlap between Superior Longitudinal Fasciculus III (SLF3) and the cortical area activated during the reading functional MRI (fMRI) task. Tractography was produced from the fully sampled (green) or the subsampled clinically-feasible diffusion MRI (purple). The mean and standard deviations of the normalized True positive rates, Dice scores, and 95% Hausdorff distances are given above the respective box whiskers. ODF-FP is compared with the best-performing reference method, i.e., FSL in the fully sampled and CSD in the subsampled data set. The p-values of the covariates (patient's age and sex, tumor hemisphere and tumor type) are given on the right side.

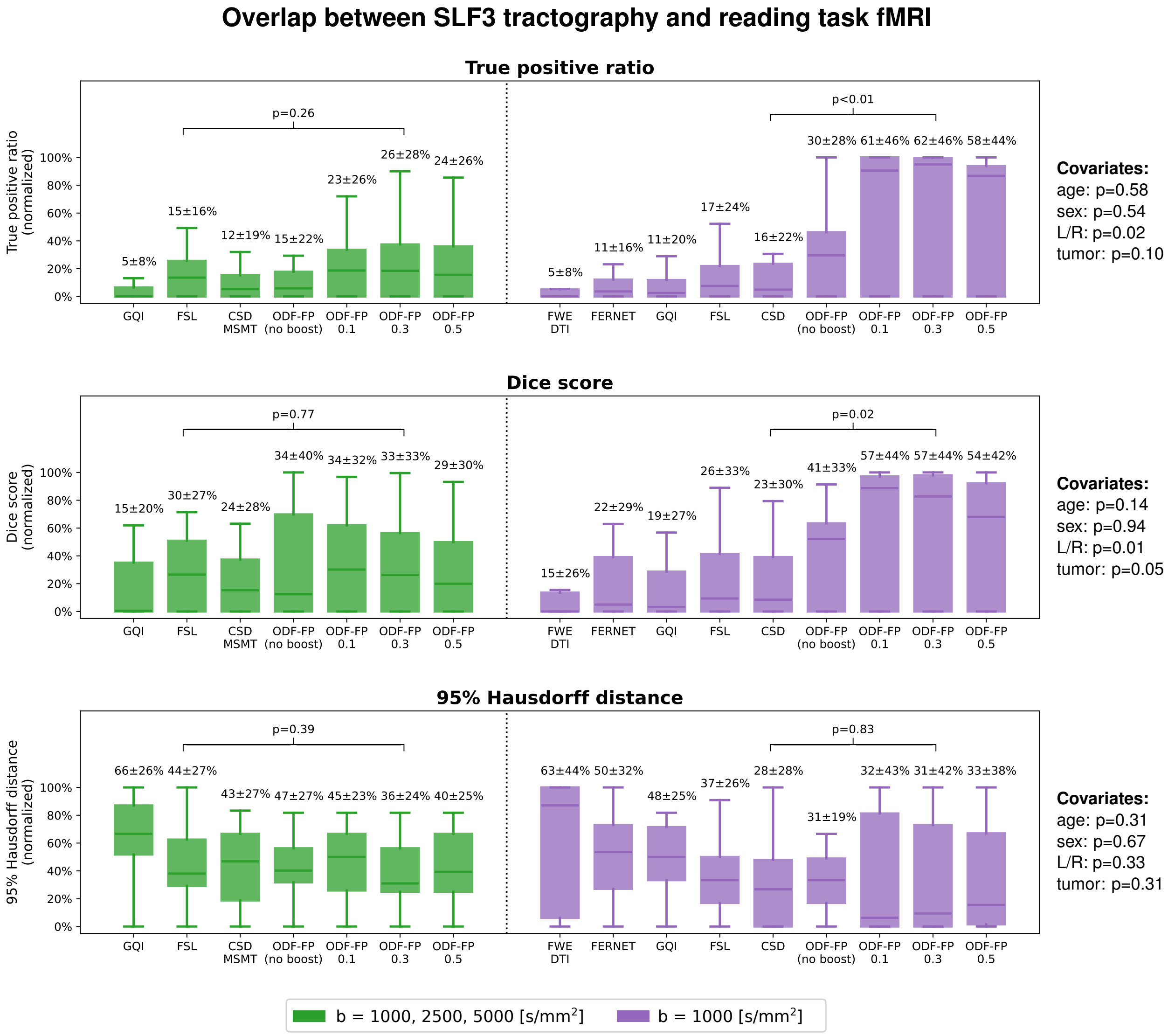


Supplementary Figure S5: Box plots of the relative overlap between Frontal Aslant Tract (FAT) and the cortical area activated during the reading functional MRI (fMRI) task. Tractography was produced from the fully sampled (green) or the subsampled clinically-feasible diffusion MRI (purple). The mean and standard deviations of the normalized True positive rates, Dice scores, and 95% Hausdorff distances are given above the respective box whiskers. ODF-FP is compared with the best-performing reference method, i.e., FSL in the fully sampled and CSD in the subsampled data set. The p-values of the covariates (patient's age and sex, tumor hemisphere and tumor type) are given on the right side.

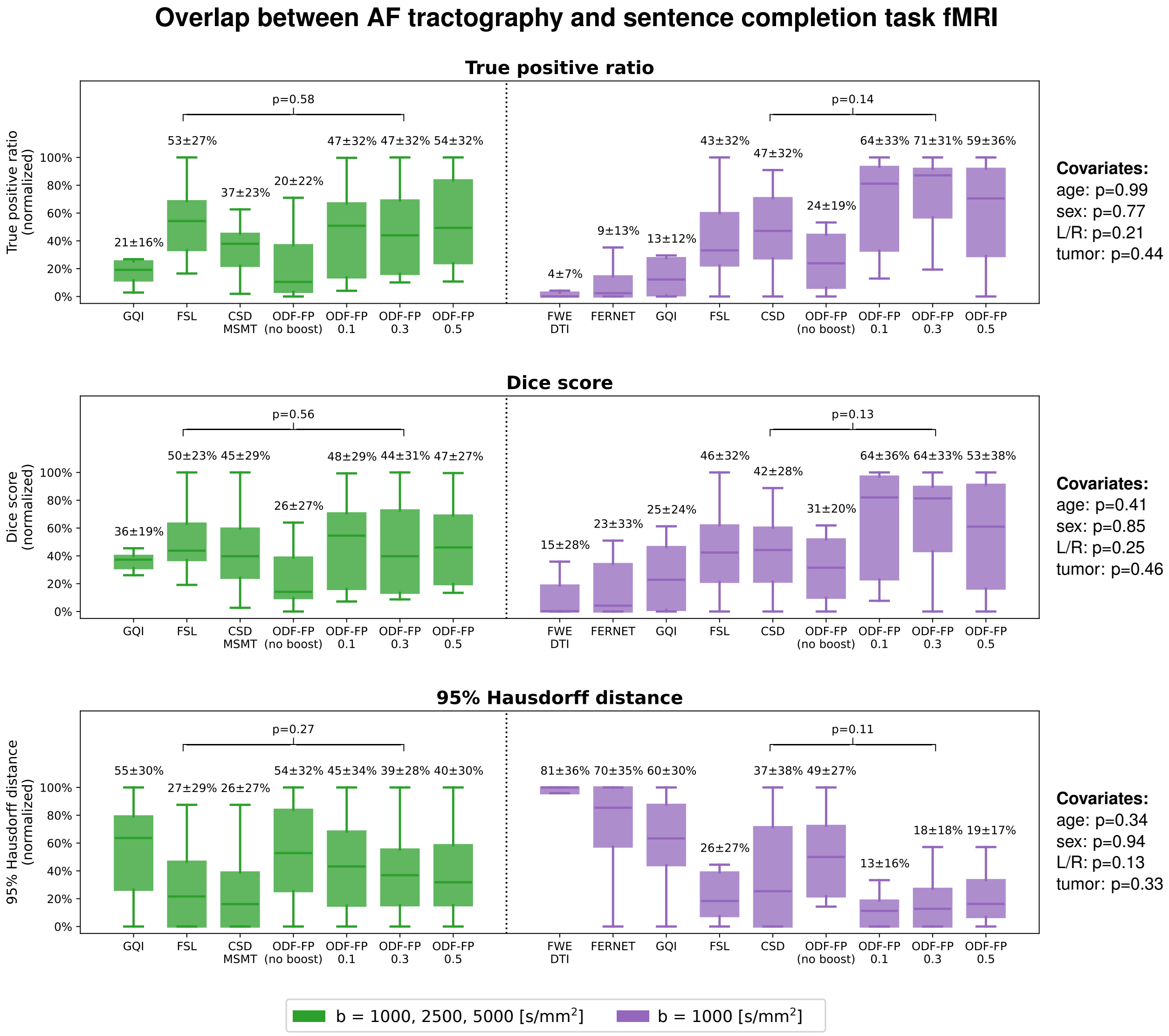


Supplementary Figure S6: Box plots of the relative overlap between Arcuate Fasciculus (AF) and the cortical area activated during the sentence completion functional MRI (fMRI) task. Tractography was produced from the fully sampled (green) or the subsampled clinically-feasible diffusion MRI (purple). The mean and standard deviations of the normalized True positive rates, Dice scores, and 95% Hausdorff distances are given above the respective box whiskers. ODF-FP is compared with the best-performing reference method, i.e., FSL in the fully sampled and CSD in the subsampled data set. The p-values of the covariates (patient's age and sex, tumor hemisphere and tumor type) are given on the right side.

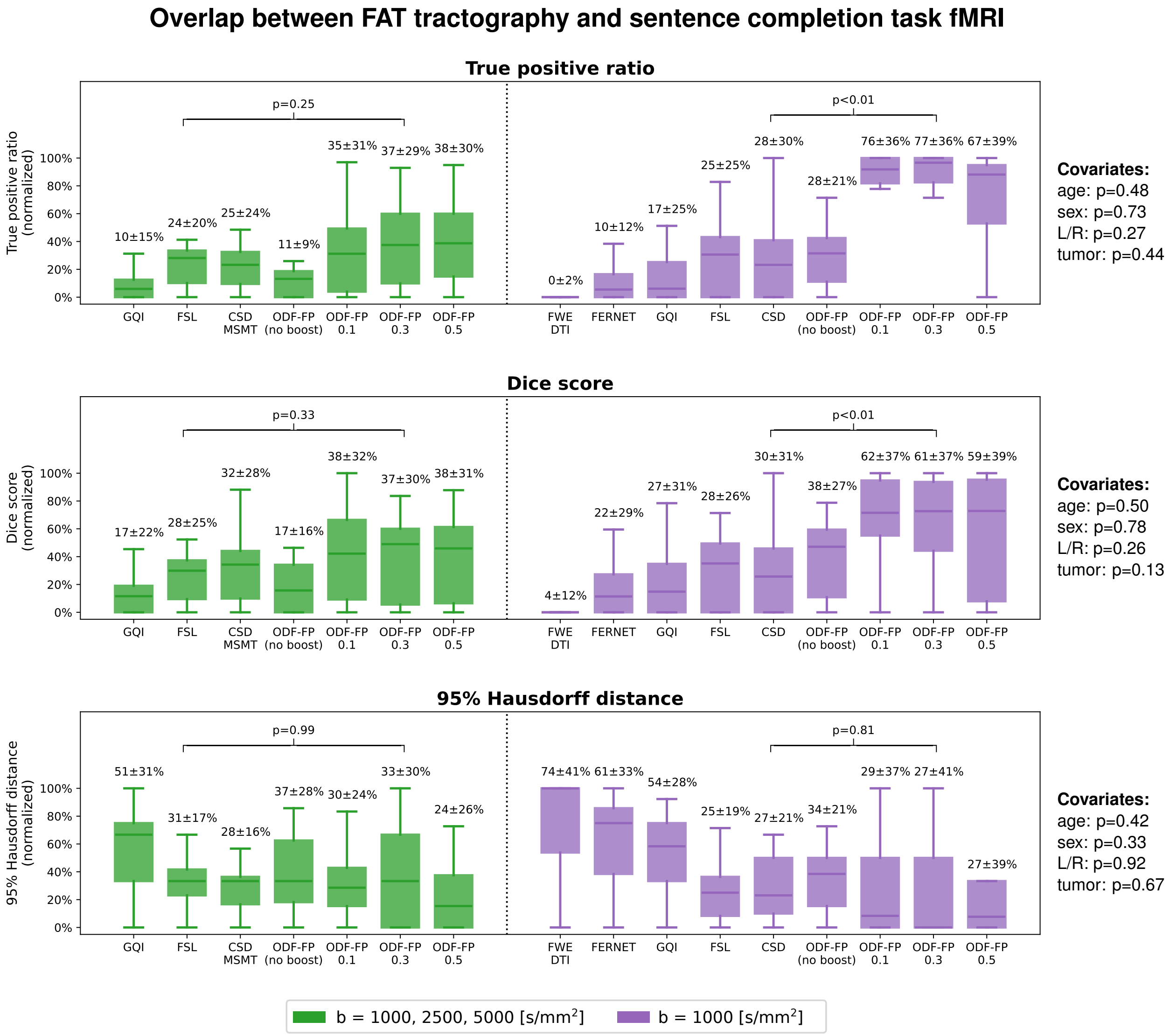


Supplementary Figure S7: Box plots of the relative overlap between Superior Longitudinal Fasciculus III (SLF3) and the cortical area activated during the sentence completion functional MRI (fMRI) task. Tractography was produced from the fully sampled (green) or the subsampled clinically-feasible diffusion MRI (purple). The mean and standard deviations of the normalized True positive rates, Dice scores, and 95% Hausdorff distances are given above the respective box whiskers. ODF-FP is compared with the best-performing reference method, i.e., FSL in the fully sampled and CSD in the subsampled data set. The p-values of the covariates (patient's age and sex, tumor hemisphere and tumor type) are given on the right side.

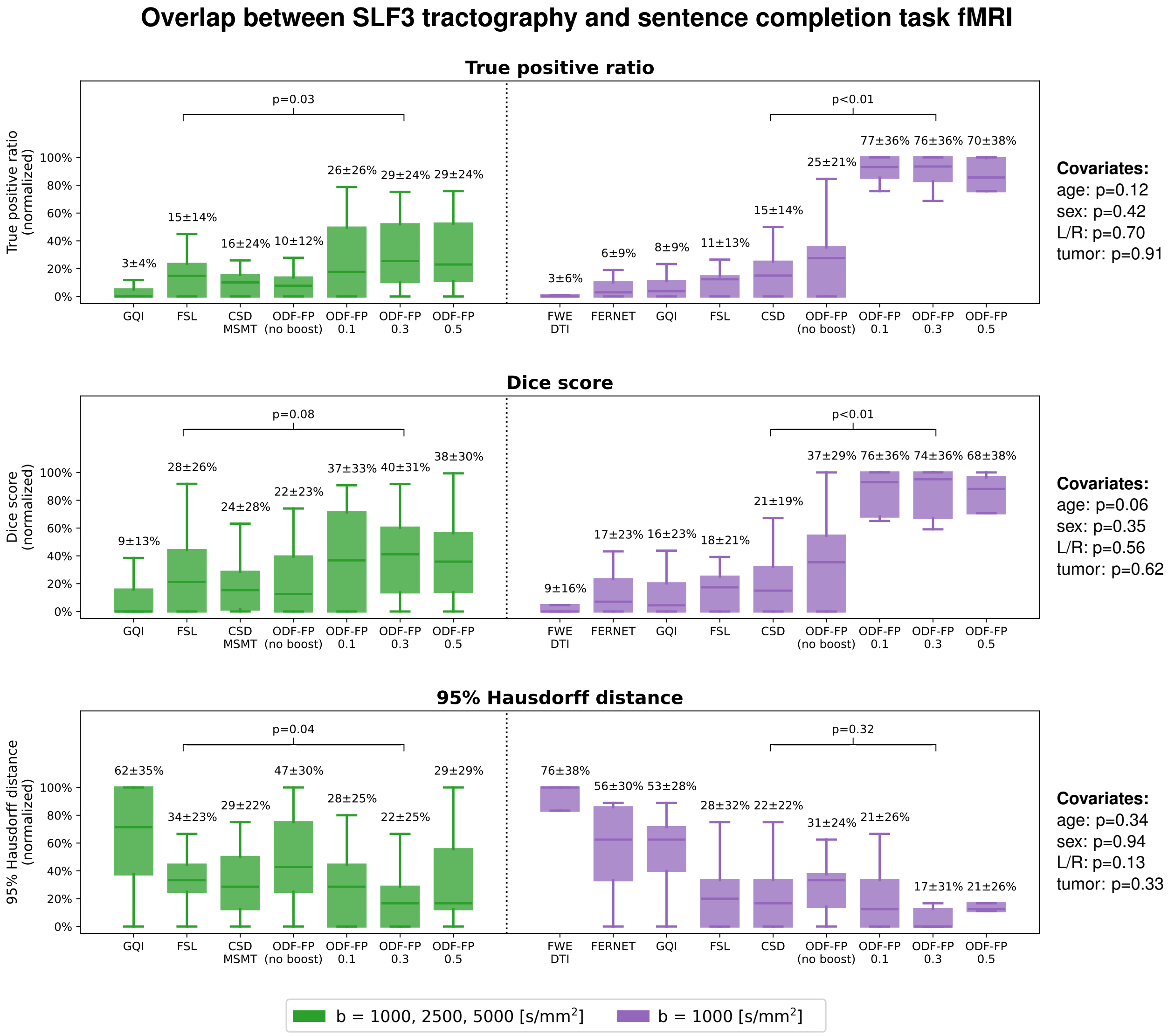


Supplementary Figure S8: Box plots of the relative overlap between Frontal Aslant Tract (FAT) and the cortical area activated during the sentence completion functional MRI (fMRI) task. Tractography was produced from the fully sampled (green) or the subsampled clinically-feasible diffusion MRI (purple). The mean and standard deviations of the normalized True positive rates, Dice scores, and 95% Hausdorff distances are given above the respective box whiskers. ODF-FP is compared with the best-performing reference method, i.e., FSL in the fully sampled and CSD in the subsampled data set. The p-values of the covariates (patient's age and sex, tumor hemisphere and tumor type) are given on the right side.

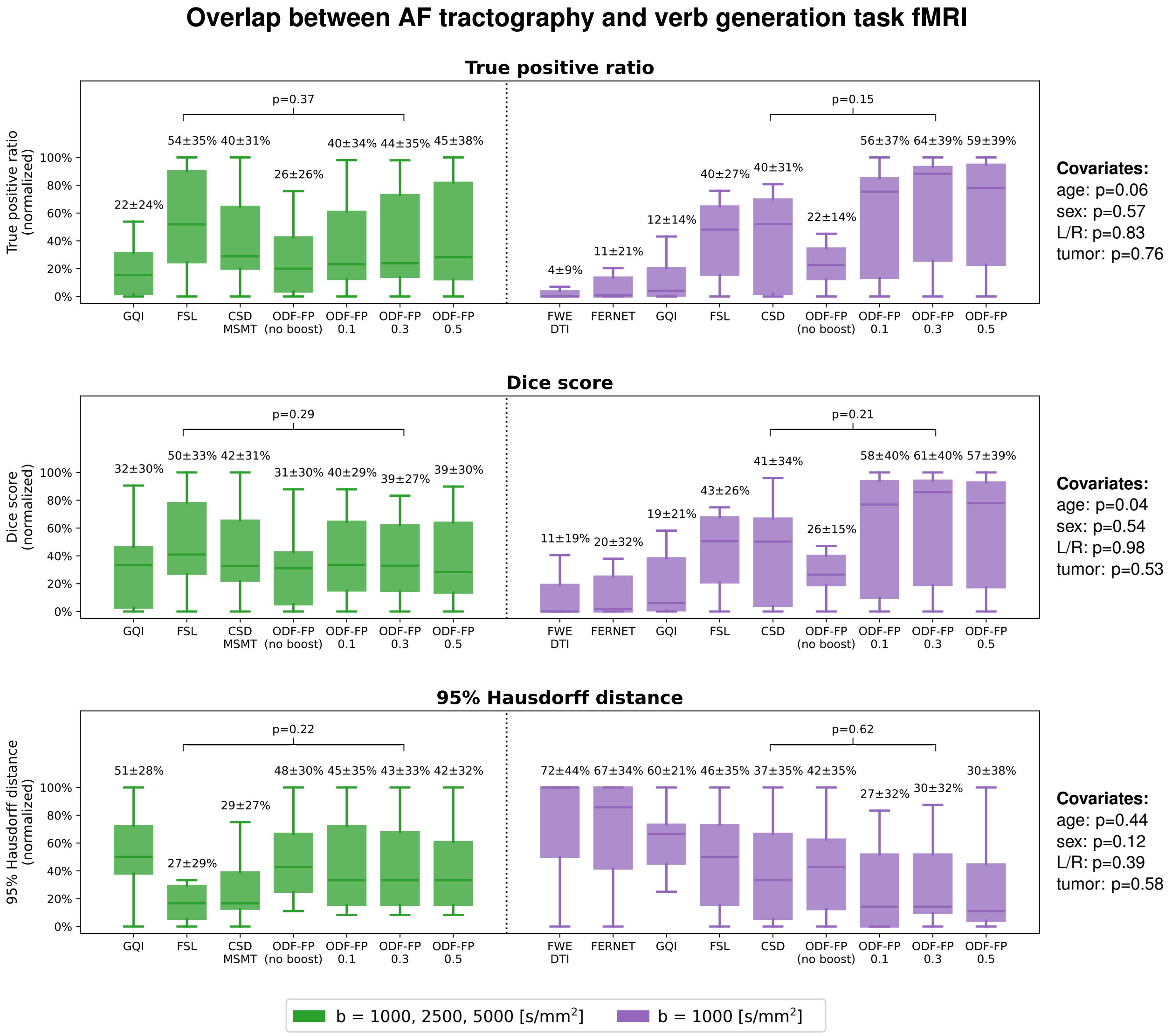


Supplementary Figure S9: Box plots of the relative overlap between Arcuate Fasciculus (AF) and the cortical area activated during the verb generation functional MRI (fMRI) task. Tractography was produced from the fully sampled (green) or the subsampled clinically-feasible diffusion MRI (purple). The mean and standard deviations of the normalized True positive rates, Dice scores, and 95% Hausdorff distances are given above the respective box whiskers. ODF-FP is compared with the best-performing reference method, i.e., FSL in the fully sampled and CSD in the subsampled data set. The p-values of the covariates (patient's age and sex, tumor hemisphere and tumor type) are given on the right side.

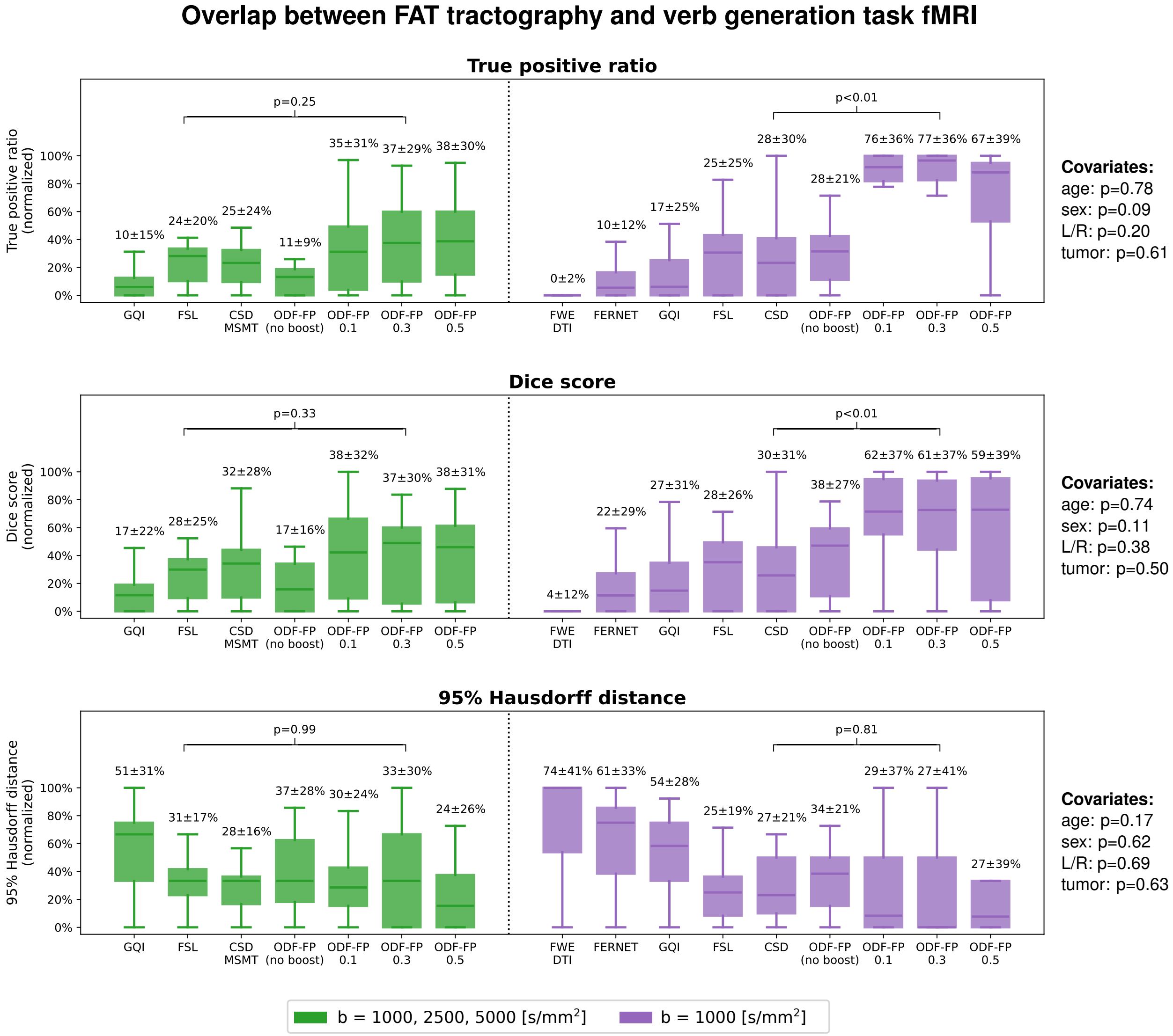


Supplementary Figure S10: Box plots of the relative overlap between Superior Longitudinal Fasciculus III (SLF3) and the cortical area activated during the verb generation functional MRI (fMRI) task. Tractography was produced from the fully sampled (green) or the subsampled clinically-feasible diffusion MRI (purple). The mean and standard deviations of the normalized True positive rates, Dice scores, and 95% Hausdorff distances are given above the respective box whiskers. ODF-FP is compared with the best-performing reference method, i.e., FSL in the fully sampled and CSD in the subsampled data set. The p-values of the covariates (patient's age and sex, tumor hemisphere and tumor type) are given on the right side.

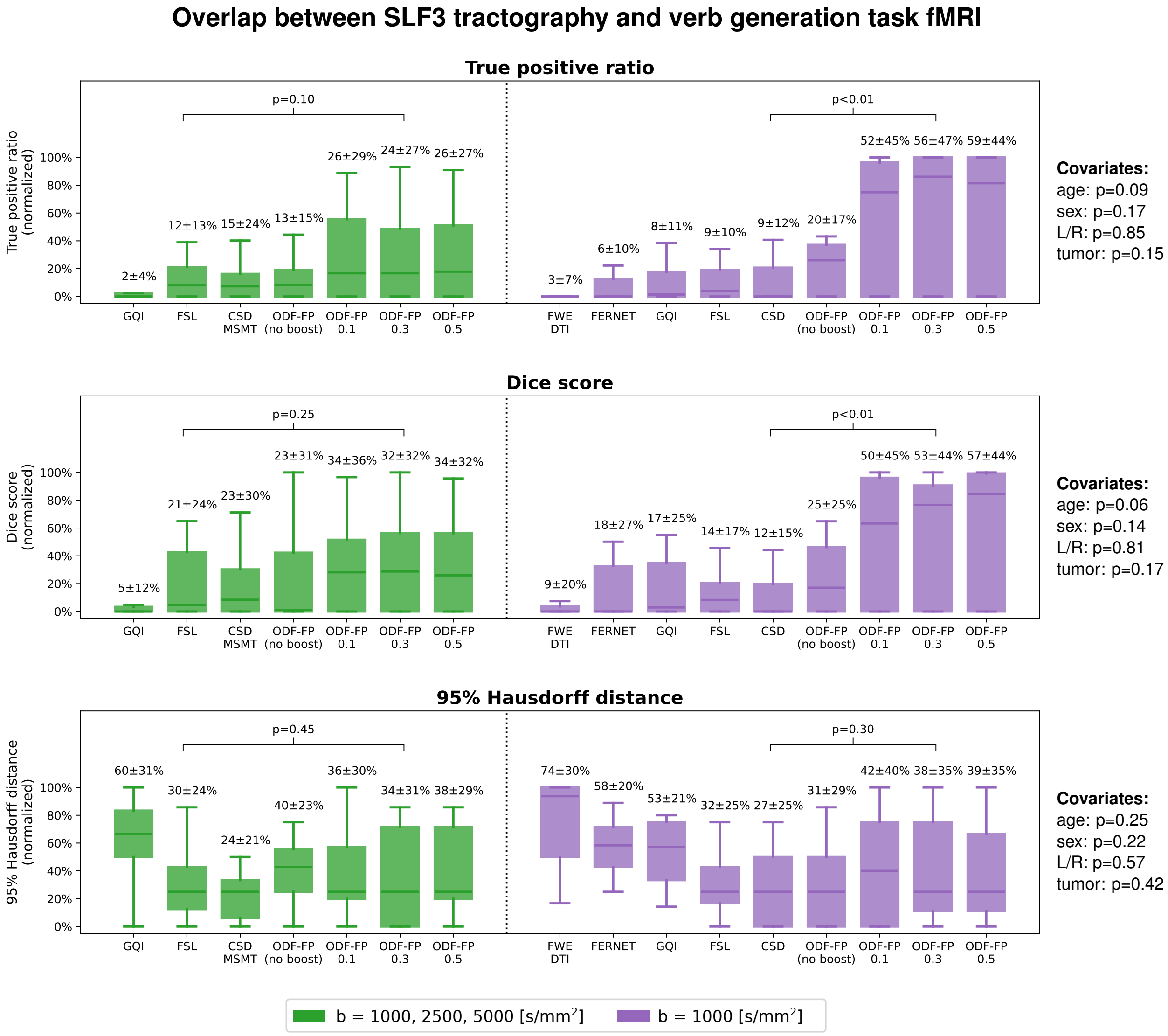


Supplementary Figure S11: Box plots of the relative overlap between Frontal Aslant Tract (FAT) and the cortical area activated during the verb generation functional MRI (fMRI) task. Tractography was produced from the fully sampled (green) or the subsampled clinically-feasible diffusion MRI (purple). The mean and standard deviations of the normalized True positive rates, Dice scores, and 95% Hausdorff distances are given above the respective box whiskers. ODF-FP is compared with the best-performing reference method, i.e., FSL in the fully sampled and CSD in the subsampled data set. The p-values of the covariates (patient's age and sex, tumor hemisphere and tumor type) are given on the right side.
